# Population analyses of mosaic X chromosome loss identify genetic drivers and widespread signatures of cellular selection

**DOI:** 10.1101/2023.01.28.23285140

**Authors:** Aoxing Liu, Giulio Genovese, Yajie Zhao, Matti Pirinen, Maryam M. Zekavat, Katherine Kentistou, Zhiyu Yang, Kai Yu, Caitlyn Vlasschaert, Xiaoxi Liu, Derek W. Brown, Georgi Hudjashov, Bryan Gorman, Joe Dennis, Weiyin Zhou, Yukihide Momozawa, Saiju Pyarajan, Vlad Tuzov, Fanny-Dhelia Pajuste, Mervi Aavikko, Timo P. Sipilä, Awaisa Ghazal, Wen-Yi Huang, Neal Freedman, Lei Song, Eugene J. Gardner, FinnGen, BCAC, MVP, Vijay G. Sankaran, Aarno Palotie, Hanna M. Ollila, Taru Tukiainen, Stephen J. Chanock, Reedik Mägi, Pradeep Natarajan, Mark J. Daly, Alexander Bick, Steven A. McCarroll, Chikashi Terao, Po-Ru Loh, Andrea Ganna, John R.B. Perry, Mitchell J. Machiela

## Abstract

Mosaic loss of the X chromosome (mLOX) is the most commonly occurring clonal somatic alteration detected in the leukocytes of women, yet little is known about its genetic determinants or phenotypic consequences. To address this, we estimated mLOX in >900,000 women across eight biobanks, identifying 10% of women with detectable X loss in approximately 2% of their leukocytes. Out of 1,253 diseases examined, women with mLOX had an elevated risk of myeloid and lymphoid leukemias and pneumonia. Genetic analyses identified 49 common variants influencing mLOX, implicating genes with established roles in chromosomal missegregation, cancer predisposition, and autoimmune diseases. Complementary exome-sequence analyses identified rare missense variants in *FBXO10* which confer a two-fold increased risk of mLOX. A small fraction of these associations were shared with mosaic Y chromosome loss in men, suggesting different biological processes drive the formation and clonal expansion of sex chromosome missegregation events. Allelic shift analyses identified alleles on the X chromosome which are preferentially retained, demonstrating that variation at many loci across the X chromosome is under cellular selection. A novel polygenic score including 44 independent X chromosome allelic shift loci correctly inferred the retained X chromosomes in 80.7% of mLOX cases in the top decile. Collectively our results support a model where germline variants predispose women to acquiring mLOX, with the allelic content of the X chromosome possibly shaping the magnitude of subsequent clonal expansion.

## Introduction

Females carry a maternal and paternal copy of the X chromosome in which one copy is partially rendered transcriptionally inactive early in development by expression of Xist^1^ and epigenetic modifications. The inactivation process is random as to which X chromosome is chosen with the resulting inactive state being irreversible and clonally transmitted to daughter cells^2^. X chromosome inactivation has evolved as a mechanism to compensate for gene dosage imbalances between XX females and XY males, although some genes are only partially inactivated^3^, including several tumor suppressor genes (e.g., *ATRX, KDM5C*)^4^. Analytic challenges associated with X inactivation and haploid male X chromosomes have led to fewer studies of the X chromosome, potentially missing critical germline and somatic variation relevant to disease risk.

With age, the expected 1:1 ratio of inactivated maternal to paternal X chromosome copies can become skewed. X chromosome inactivation skewing is observed in various tissues with high frequencies observed in leukocytes^5,6^. Detectable skewed X chromosome inactivation in leukocytes is heritable (h^2^=0.34)^7^ and can indicate depletion of haematopoietic stem cells, selection pressures on leukocytes, or clonal hematopoiesis (CH). Recent investigations of age-related CH have described elevated rates of mosaic sex chromosome aneuploidies in population-based surveys of apparently healthy adults^8-13^. Mosaic loss of the female X chromosome (mLOX) is elevated in frequency compared to the autosomes^14^, preferentially impacts the inactivated X chromosome^10^ and is associated with elevated leukemia risk^15,16^. This contrasts with the male X chromosome which has very low rates of aneuploidy^17^. As the X chromosome encompasses approximately 5% of the genome and contains genes relevant to immunity, cancer susceptibility, and cardiovascular diseases, loss of a homologous copy and subsequent hemizygous selection could lead to downstream consequences on female health, as observed in Turner syndrome (45,XO)^18^; however, no study has systematically examined longitudinal associations of mLOX with disease risk.

As mLOX is a clonal pro-proliferative genomic alteration, understanding the molecular mechanisms driving susceptibility to mLOX could provide new insights into the impact of aging on hematopoiesis as well as hematologic cancer risk. The X chromosome, particularly the inactive X, is more frequently mutated in cancer genomes^19^ and is late replicating relative to autosomes, potentially increasing susceptibility to chromosomal alterations^20^. While few genome-wide association studies (GWAS) of mLOX have been reported to date^14,21^, GWAS of mosaic loss of the Y chromosome (mLOY) in men has identified hundreds of susceptibility loci^11-13,22^, many of which highlight genes involved in cell cycle regulation and cancer susceptibility. Here we describe insights from epidemiologic and genetic analyses of X chromosome loss from a combined meta-analysis of 904,524 women. We identify 49 independent common susceptibility variants across 35 loci, rare missense variants of *FBXO10* associated with mLOX, and 44 X chromosome loci that strongly influence which X chromosome is retained. The identified signals only partially overlap with known signals for other age-related types of CH. These data indicate mLOX, along with other age-related types of CH, are important pre-clinical indicators of hematologic cancer risk and identify mitotic missegregation, autoimmunity, blood cell trait, and cancer predisposition genes as core etiologic components for mLOX susceptibility and selection.

## Results

### Mosaic loss of the X chromosome in eight contributed biobanks

We leveraged genetic data in a total of 904,524 women from eight biobanks worldwide, including European ancestry participants from FinnGen^23^, Estonian Biobank (EBB)^24^, UK Biobank (UKBB)^25,26^, Breast Cancer Association Consortium (BCAC)^27,28^, Million Veteran Program (MVP)^29,30^, Mass General Brigham Biobank (MGB)^31,32^, and Prostate, Lung, Colorectal and Ovarian Cancer Screening Trial (PLCO)^33^, as well as East Asian ancestry participants from Biobank Japan (BBJ)^34^ (**Supplementary Table S1**). The median (SD) age at sample collection for genotyping ranged from 44 (16.3) for EBB to 67.2 (12.9) for FinnGen. We identified mLOX using the Mosaic Chromosomal Alterations (MoChA) WDL pipeline (https://github.com/freeseek/mochawdl), which uses raw signal intensities from single-nucleotide polymorphism (SNP) array data. Out of 904,524 women, 86,093 (9.5%) were classified as cases with detectable mLOX (**Methods**; **Table 1**). Overall, the cell fraction of mLOX (i.e., the estimated fraction of peripheral leukocytes with X loss) was low (median=2.0%) with expanded clones having frequency ≥5% infrequently observed (0.5% of women) (**Supplementary Figure S1**). A subset of UKBB participants (243,520 out of 261,145) also had whole-exome sequencing (WES) data available which allowed us to assess the performance of mLOX calling from MoChA. Among UKBB mLOX cases classified by MoChA, a high correlation (r=-0.86) was observed between cell fraction derived from SNP array data (by MoChA) and X dosage derived from WES data (**Supplementary Figure S2**). In addition to the MoChA generated dichotomous measure used by all biobanks, in UKBB we generated a 3-way combined quantitative measure by integrating independent information from both SNP array and WES data (**Methods**). The t-test statistic for association with age was increased by 29.2% with the 3-way calls, indicating improved performance relative to SNP array-only calls.

**Table 1.** Descriptive characteristics of the eight biobanks contributing to the mLOX analysis.

| Biobank | Median age (SD) | mLOX Cases | Controls | Effective sample size | Continental ancestry groups |
| --- | --- | --- | --- | --- | --- |
| FinnGen | 67.2 (12.9) | 27,001 | 141,837 | 90,732 | European, Finnish |
| Estonian Biobank (EBB) | 44 (16.3) | 20,232 | 110,547 | 68,408 | European, Estonians |
| UK Biobank (UKBB) | 57 (8.0) | 16,214 | 244,931 | 60,829 | European, British |
| Biobank Japan (BBJ) | 65 (15.8) | 13,597 | 63,720 | 44,823 | East Asian, Japanese |
| Breast Cancer Association Consortium (BCAC) | 57 (11.3) | 2,773 | 195,499 | 10,937 | European |
| Million Veteran Program (MVP) | 54 (13.9) | 1,496 | 33,192 | 5,726 | European |
| Mass General Brigham Biobank (MGB) | 54 (17.3) | 2,108 | 11,527 | 7,128 | European |
| Prostate, Lung, Colorectal and Ovarian Cancer Screening Trial (PLCO) | 64.0 (5.4) | 2,672 | 17,178 | 9,249 | European |

### Environmental determinants and epidemiological consequences

Like many other types of somatic mutations^13,14^, the frequency of women with detectable mLOX in peripheral leukocyte is age-related, with a frequency of 2.5% in women aged <40 and reaching >32.2% after 80, averaged over all contributing biobanks (**Supplementary Figure S3** and **Table S2**). To investigate the effect of lifestyle factors on the risk of acquiring detectable mLOX, we assessed associations of smoking and body mass index (BMI) with mLOX in the FinnGen cohort, which had an available smoking status for 50.3% of women and BMI for 18.4% of women (**Methods**). Overall, ever-smokers had no increased risk of mLOX (P=0.56); however, an increased risk was observed among ever-smokers for acquiring expanded mLOX with cell fraction ≥5% (OR=1.3 [1.2-1.5], P=6.9×10^−5^) (**Supplementary Table S3** and **Figure S4-5**). The relationship between smoking and skewed X-inactivation has not been established, as smoking was suggested as a modulator for skewed XCI in the whole-blood tissue for women older than age 55^7^ but not associated in the TwinsUK cohort^35^. No associations were observed between BMI and mLOX (**Supplementary Table S4**).

To evaluate disease outcomes associated with detectable mLOX, we performed Cox proportional hazards regression for incident disease cases in FinnGen, UKBB, MVP, and MGB independently considering genotyping age and ever-smoking status as covariates and meta-analyzed across biobanks with a fixed-effect model (**Methods**). Out of the 1,253 diseases we examined, we identified mLOX associations (P<4.0×10^−5^) with leukemia overall (HR=1.7 [1.5-2.1], P=3.5×10^−10^) and chronic lymphoid leukemia (HR=3.3 [2.4-4.4], P=8.4×10^−15^) and suggestive evidence for acute myeloid leukemia (AML) (HR=1.9 [1.3-2.8], P=1.8×10^−3^) (**Supplementary Table S5**). Unlike the germline loss of the X chromosome in women with Turner syndrome (45,XO), which can cause various medical and developmental problems^18^, we noted limited clinical consequences for women with detectable mLOX in blood.

As the average mLOX cell fraction impacted is approximately 2%, we proposed that investigating expanded mLOX with higher cell fraction (≥10% as previously defined^16^) could result in stronger disease associations. Restricting to expanded mLOX, we observed evidence for elevated associations with leukemia overall (HR=6.3 [3.9-10.2], P=7.3×10^−14^) and AML (HR=10.6 [3.1-36.1], P=1.5×10^−4^) (**Supplementary Table S6**). We also observed suggestive evidence for associations with vitamin B complex deficiency (HR=3.7 [1.8-7.9], P=6.0×10^−4^) and pneumonia (HR=1.5 [1.2-1.8], P=4.7×10^−4^), especially pneumonia caused by bacterial infections (HR=1.8 [1.3-2.3], P=3.9×10^−5^). Similarly, in UKBB^16^, an increased risk of incident pneumonia was observed for both women with expanded mLOX (HR=1.8 [1.0-3.2], P=0.035) and men with expanded mLOY (HR=1.2 [1.1-1.4], P=1.1×10^−4^).

To examine the potential impacts of other types of CH on mLOX associations with leukemia, we performed sensitivity analyses in UKBB where we had available calls on autosomal mosaic chromosomal alterations (mCAs) as well as CH mutations in driver genes, commonly referred to as clonal hematopoiesis of indeterminate potential (CHIP)^36^. We observed attenuations in associations for expanded mLOX when removing individuals with autosomal mCAs (HR=3.8 [1.6-9.3], P=2.7×10^−3^), CHIP (HR=6.2 [3.1-12.4], P=3.1×10^−7^) and both mCAs and CHIP (HR=4.5 [1.9-10.8], P=8.6×10^−4^) (**Supplementary Table S7**); however, significant associations with expanded mLOX and overall leukemia risk remained indicating mLOX is independently associated with leukemia risk. Associations for other lymphoid and myeloid leukemias display similar patterns, albeit losing statistical significance likely due to reduced sample size (**Supplementary Table S7**).

### Common and rare variants associated with mLOX susceptibility

We performed a genome-wide association study (GWAS) to identify common and low-frequency germline variants (minor allele frequency (MAF)>0.1%) associated with the risk of developing detectable mLOX in peripheral leukocytes. We examined the autosomes (chromosomes 1-22) and X chromosome in each of the eight contributing biobanks independently, for a total of 904,524 women (**Methods**). To increase GWAS power, we used enhanced 3-way combined calls for UKBB and meta-analyzed summary statistics across different mLOX measures with a weighted z-score method (**Methods**). Of the 33,737,466 variants examined, we identified 49 independent genome-wide significant variants (P<5.0×10^−8^) across 35 loci associated with mLOX susceptibility (**Methods**; **Figure 2A**; **Supplementary Table S8**). Most independent variants were located on chromosome 6 (21 variants), 2 (7 variants), 17 (3 variants), and X (3 variants). Despite differences in age-adjusted mLOX frequencies, mLOX variant effects were consistent across the eight biobanks and across European and East Asian ancestry (P from Cochran’s Q-test <0.05/49 = 0.001) (**Supplementary Table S9**), with the exception of rs9267499 (*HSPA1A*, P from meta-analysis = 1.4×10^−21^, P from heterogeneity test=5.8×10^−4^) and rs78378222 (*TP53*, P from meta-analysis = 3.3×10^−10^, P from heterogeneity test = 7.1×10^−4^). For rs9267499, the association was predominantly driven by FinnGen (P from GWAS = 1.3×10^−20^), for which the risk allele C had higher frequency (16%) in this Finnish European population compared with other biobanks with either non-Finnish European (8-10%) or East Asian (3%) ancestry. For rs78378222, the heterogeneity of variant effects across biobanks was likely due to differences in mLOX cell fraction by contributing studies. When stratifying by cell fraction in FinnGen, the OR for the risk allele of rs78378222 was 1.12 [1.03-1.21] (P=0.01) for cell fractions below 5% but reached 1.73 [1.30-2.29] (P=1.4×10^−4^) for expanded mLOX with cell fraction above 5% (P for effect size difference from a two-sided t-test = 2.5×10^−5^) (**Supplementary Table S10** and **Figure S7**).

**Figure 1.**
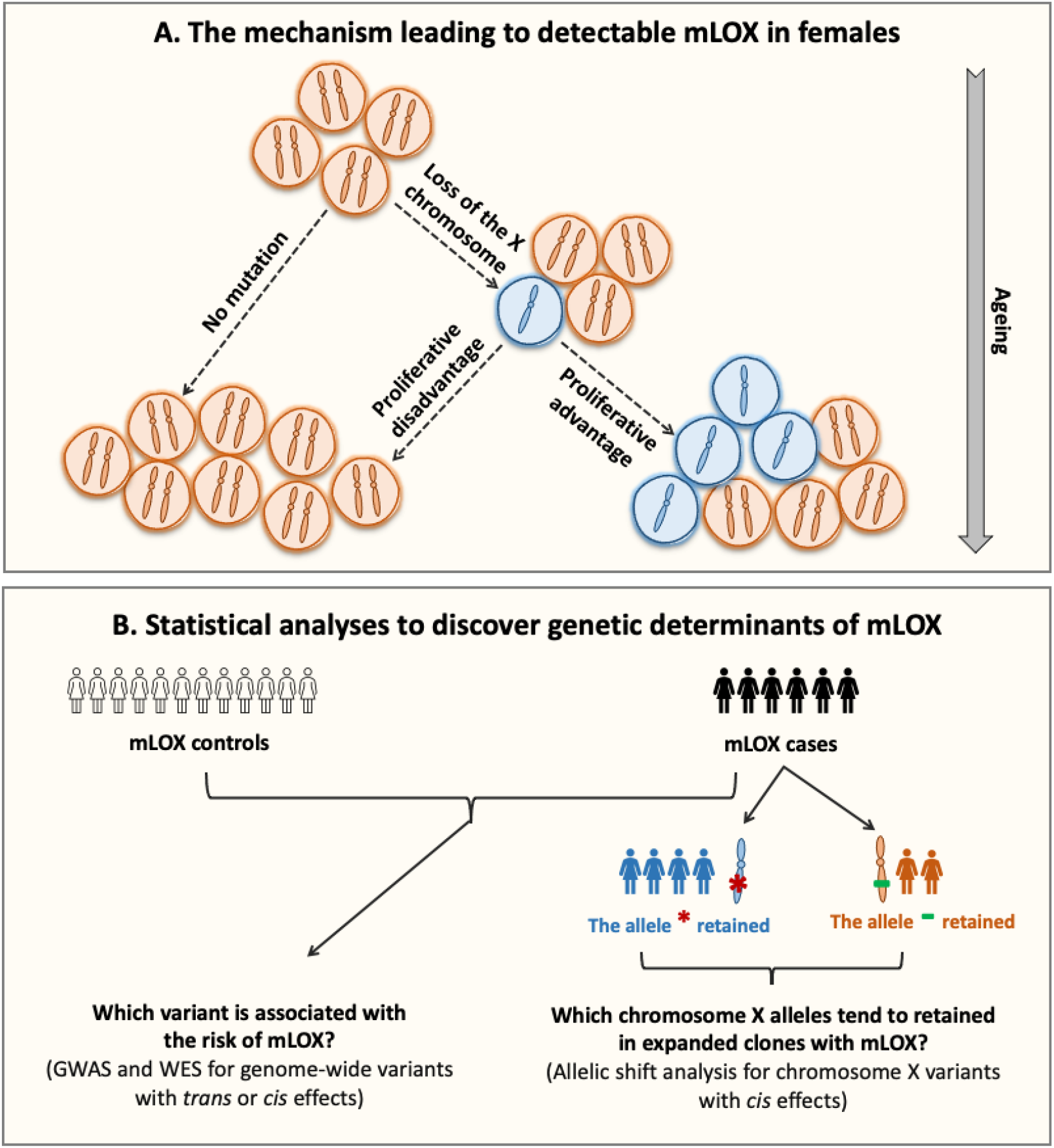
Theoretical framework of the mLOX study. Panel **(A)** depicts the etiologic process leading to detectable mosaic loss of the X chromosome (mLOX) in females. Detectable age-related mLOX develops only if the mutant haematopoietic stem cell (HSC) survives loss of the X chromosome and the mutation confers a proliferative advantage over normal cells. Panel **(B)** shows the statistical approaches used to discover the genetic determinants of mLOX. Variants associated with susceptibility to mLOX, acting as either *trans* or *cis* factors, are examined using a genome-wide association study (GWAS), for common variants with minor allele frequency (MAF) > 0.1%, and a gene-burden test performed for whole-exome sequencing (WES) data for rare variants with MAF < 0.1%. Among samples with detectable mLOX, allelic shift analysis is used to detect chromosome X alleles exhibiting *cis* selection, that is, more likely to be clonally selected for when detectable mLOX retains these alleles.

**Figure 2.**
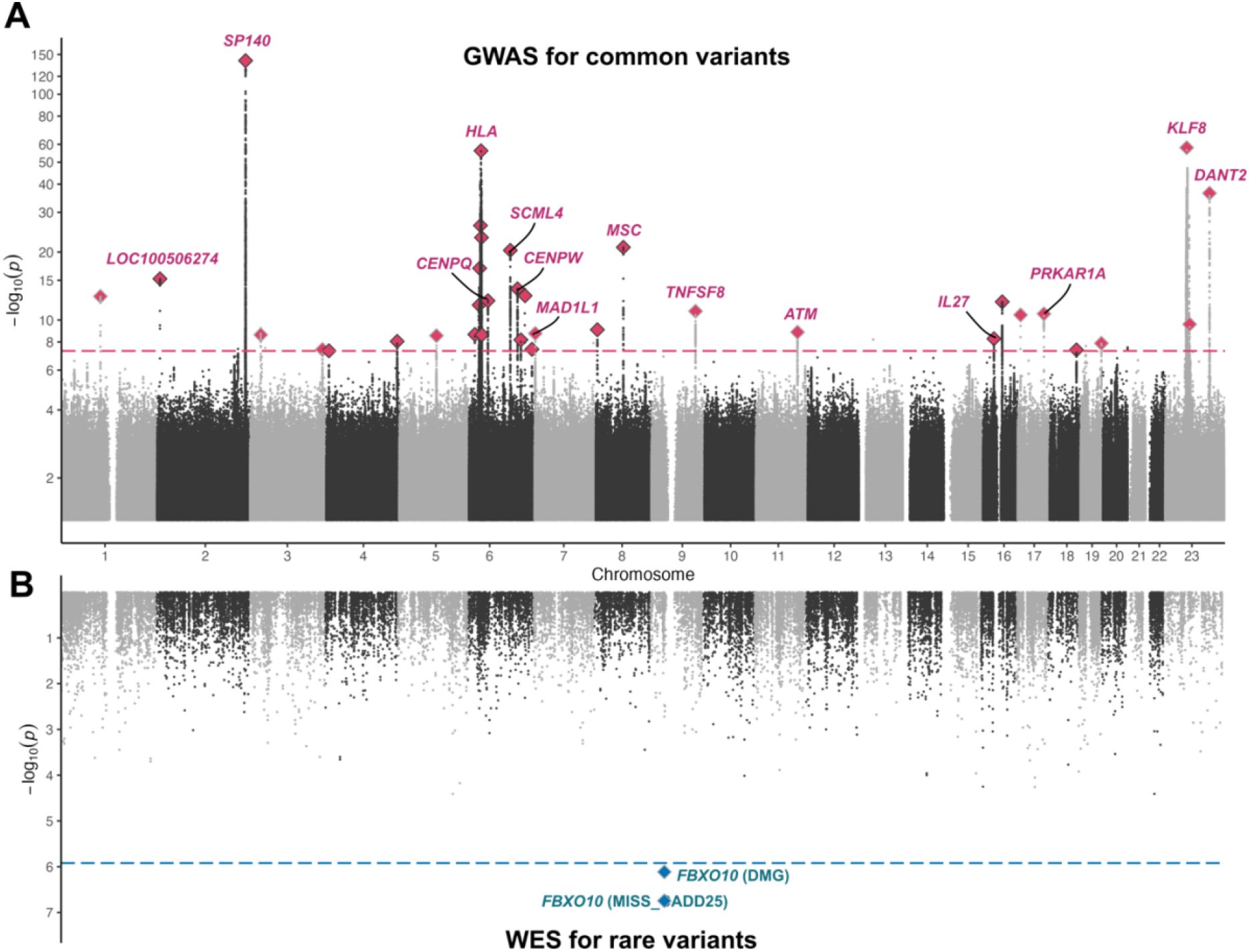
Common and rare genetic contributors to mLOX susceptibility. Panel (**A**) shows genome-wide association study -log_10_(P) for the association of common variants (MAF>0.1%) with mLOX. Labels are only assigned for candidate genes of the top 10 lead variants from meta-analysis or the top 10 candidate genes from gene prioritization and the y-axis is log scale. Panel (**B**) presents gene burden test -log_10_(P) for the rare variants (MAF<0.1%) associations with mLOX. The dashed lines denote the statistical significance, which is 5.0×10^−8^ for GWAS (**A**) and 1.2×10^−6^ for the gene-burden test (**B**).

We deployed a range of variant to gene mapping approaches to rank genes proximal to each of our hits by their strength of evidence for causality (**Methods**), highlighting the highest-scoring gene at each locus (**Supplementary Table S11**). The most significantly associated mLOX locus is at 2q37.1 which we mapped to *SP140*, an interferon-inducible gene expressed at high levels in leukocytes with nearby genetic variants associated with chronic lymphocytic leukemia^37^ and autoimmune diseases^38,39^. Several identified mLOX loci implicated plausible causal genes relevant to cancer predisposition including *JARID2* (6p22.3), *MYB* (6q23.3), *TNFSF8* (9q32-q33.1), *ATM* (11q22.3), *TP53* (17p13.1), *PRKAR1A* (17q24.2), and *KLF8* (Xp11.21), many of which (e.g., *JARID2*^40^, *MYB*^41^, *ATM*^42^, *TP53*^43^, and *PRKAR1A*^44^) are directly relevant to leukemia predisposition or progression. Additionally, highlighted genes at several mLOX loci are important for mitotic spindle assembly and kinetochore function including *MAD1L1* (7p22.3), *CENPU* (4q35.1), *CENPQ* (6p12.3), and *CENPW* (6q22.32), all of which are highly relevant to mitotic missegregation errors leading to loss of an X chromosome at a single cell level. Several mLOX associated loci also implicate genes related to immunity and autoimmune disorders including *EOMES* (3p24.1), *ERAP2* (5q15), *HLA-A* (6p22.1), *HLA-B* (6p21.33), *AGER* (6p21.32), *HLA-DPA1* (6p21.32), *IL27* (16p12.1-p11.2), and *LILRA1* (19q13.42), suggesting a shared etiologic relationship between mLOX and immune cell function. Similar to these locus-specific results, the genome-wide pathway-based analysis identified enrichment in pathways related to DNA damage response, cell-cycle regulation, cancer susceptibility, and immunity (**Methods**; **Supplementary Table S12**).

We next investigated if the identified common variants for mLOX susceptibility in women were associated with mLOY, the most common leukocyte sex chromosome mosaicism in men (**Supplementary Figure S8**) and likewise if mLOY loci were associated with mLOX. We employed a Bayesian model to assign 49 independent common variants identified from mLOX GWAS and 147 variants (nine variants dropped due to missing in mLOX GWAS) from the published mLOY GWAS^13^ into three groups: specific to mLOX, specific to mLOY, and shared between mLOX and mLOY (**Methods; Figure 3A**). Out of 49 variants identified from the mLOX GWAS, we assigned 36 variants as specific for mLOX and eight as shared with mLOY, with greater than 95% probability (**Supplementary Table S13**). Among three centromere protein genes identified for mLOX susceptibility, *CENPQ* (for rs2448705, OR=0.96 [0.95-0.97] for mLOX and 1.00 [0.99-1.02] for mLOY, P for effect size difference=6.16×10^−8^) and *CENPW* (for rs9398805, OR=1.04 [1.03-1.06] for mLOX and 1.02 [1.01-1.04] for mLOY, P for effect size difference=0.01) were specific to mLOX with posterior probability > 95%, while for *CENPU* (for rs2705883, OR=1.04 [1.03-1.06] for mLOX and 1.03 [1.01-1.04] for mLOY, P for effect size difference=0.09) the probability to be mLOX specific was 91%. When likewise examining the 147 mLOY susceptibility variants, we further identified nine variants (prioritized genes such as *SPDL1, HLA-A, CHEK2*, and *MAGEH1*) to be shared with mLOX susceptibility, in addition to the six variants that are exactly mLOX GWAS lead variants (prioritized genes *GRPEL1, QKI, TP53*, and *MAD1L1*) or in high LD (r^2^>0.6) with mLOX GWAS lead variants (prioritized genes *ATM* and *HEATR3*). Notably, for variants that are shared between mLOX and mLOY, ORs were attenuated for mLOX relative to mLOY, possibly due to lower cell fractions observed for mLOX as compared to mLOY (**Supplementary Figure S1**). For example, for rs78378222 (*TP53*), the effect size for mLOX (OR=1.16 [1.11-1.22]) was lower than for mLOY (OR=1.77 [1.65-1.88]) (P for effect size difference=3.25×10^−10^). Likewise for rs2280548 (*MAD1L1*), the effect for mLOX (OR=1.03 [1.02-1.05]) was also lower than for mLOY (OR=1.13 [1.11-1.14]) (P for effect size difference=9.13×10^−9^). This smaller effect size together with the lower frequency of mLOX (e.g., 6.2% for 261,145 women in UKBB aged 40-70 at genotyping) relative to mLOY (e.g., 20.4% for 205,011 men in UKBB aged 40-70 at genotyping^13^) indicates that a large meta-analysis was needed to identify susceptibility variants for mLOX. The partially shared genetic architecture from common variants between mLOX and mLOY was also supported by the moderate genetic correlation (r=0.30 [0.20-0.40], P=2.9×10^−9^) (**Methods; Supplementary Table S14**). We note that, in addition to potential differences in biological mechanisms, the differences between mLOX and mLOY could also be related to differences in cell fractions as calling algorithms can detect lower cell fraction mLOX events relative to mLOY events (**Supplementary Figure S1**).

**Figure 3.**
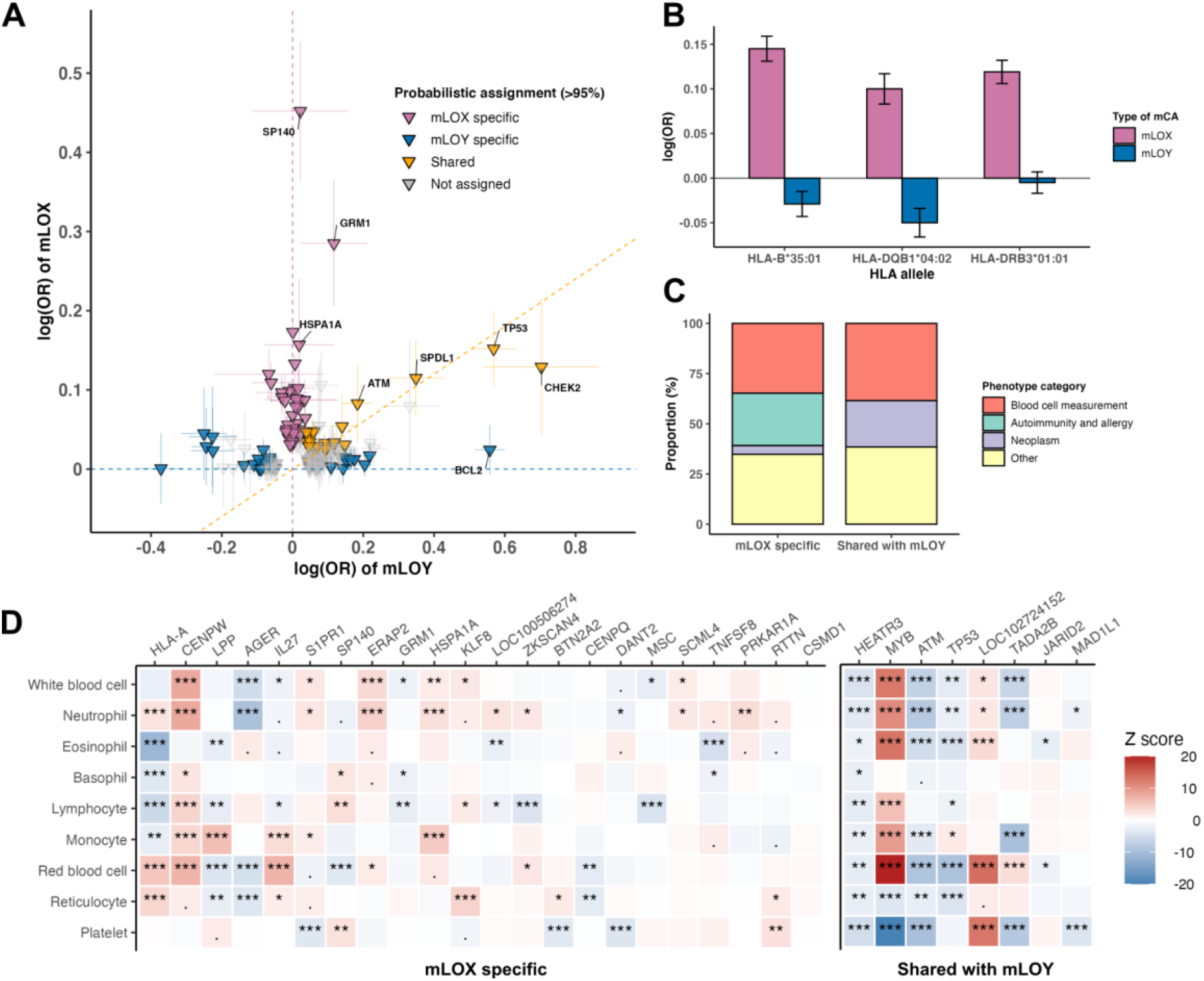
Shared and distinct genetic contributors to mLOX susceptibility in women and mLOY susceptibility in men. Examination of the shared and distinct genetic contributors to mLOX in women and mLOY in men. Panel (**A**) is a scatterplot of mLOX susceptibility variants (N=49) and mLOY susceptibility variants^13^ (N=147) and their effects on mLOX and mLOY. Variants are assigned to mLOX specific, mLOY specific, and shared by applying a Bayesian model with posterior probability >95%. (**B**) Fine-mapping of imputed HLA alleles for mLOX and mLOY in FinnGen, for three HLA alleles that are significantly associated with mLOX from step-wise conditional analyses. Panel (**C**) and (**D**) depict phenotype associations for lead variants of 30 independent mLOX susceptibility loci that were assigned to either mLOX specific or shared with mLOY. (**C**) Phenotype associations (GWAS lead variants (r^2^>0.6)) from Open Targets genetics. To avoid the impact of pleiotropic effects, we categorized phenotypes into blood cell measurement, autoimmunity and allergy, neoplasm, and others. The association with each phenotype category was first examined at a variant level and then summarized over all variants assigned to the same category in terms of the relationship with mLOY. To avoid the associations driven by HLA signals, we excluded all identified variants from the extended MHC region (GRCh38: chr6:25.7-33.4 Mb). (**D**) Associations with nine blood cell count traits^47^. The absolute Z scores were cropped to the range of [0-20].

Given the many associations of HLA genes with mLOX, we fine-mapped HLA alleles at a unique protein sequence level on 10 genes commonly used for HLA marker matching in organ transplantation for a set of 168,838 Finnish female participants (N of mLOX cases=27,001) and 128,729 Finnish male participants (N of mLOY cases=45,675) (**Methods**). Out of 156 examined HLA alleles, 16 alleles were associated with the odds of developing detectable mLOX (P<5.0×10^−8^), including alleles from both MHC class I (6 out of 74 examined alleles locating on HLA-A, -B, and - C) and class II molecules (10 out of 82 examined alleles locating on HLA-DR, -DP, and -DQ) (**Supplementary Table S15**). The most significant HLA allele HLA-B*35:01 increased the risk of mLOX (OR=1.16 [1.12-1.19], P=1.1×10^−23^), but had no effect on mLOY (OR=0.97 [0.94-1.00], P for mLOY=0.03, P for effect difference with mLOX = 3.6×10^−18^) (**Figure 3B**). This association with HLA-B*35:01 was independently replicated in BBJ (OR= 1.10 [1.05-1.15], P=1.5×10^−5^). The HLA-B*35:01 allele is well established as the major driver for the progression of human immunodeficiency virus (HIV)^45^ and also associated with several autoimmune diseases (e.g., subacute thyroiditis (OR=4.36 [3.25-5.85])^46^). With stepwise conditional analyses in FinnGen, we identified two independent genome-wide significant HLA associations at HLA-DRB3*01:01 (copy number variation that presents only in a subset of individuals) (OR=0.89 [0.87-0.91], P=2.8×10^−19^) and HLA-DQB1*04:02 (OR=0.90 [0.87-0.94], P=6.5×10^−9^). For mLOY in males, despite a larger effective sample size, no HLA allele reached the genome-wide significant threshold suggesting that HLA has a larger role in mLOX than mLOY. Additionally, we conducted conditional GWAS analyses in FinnGen by adjusting for the three lead variants (rs74615740 (*HLA-B*) (r2=0.45 with HLA-B*35:01), rs9275511 (*HLA-DQA2*), rs2734971 (*HLA-G*)) identified from the Finnish population GWAS. The results suggest that the associations with mLOX observed in the extended MHC region (GRCh38: chr6:25.7-33.4 Mb) were likely due to HLA signals instead of nearby non-HLA variants (**Supplementary Figure S9**).

To understand potential mechanisms relevant to mLOX susceptibility revealed by each identified mLOX variant, we examined associations with additional phenotypes documented in the Open Target Genetics platform (**Methods**). Out of 49 independent variants, 26 were in LD (r^2^>0.6) with at least one GWAS lead variant from Open Target (5.0×10^−8^) (**Supplementary Table S16**). Notably, more than half of the phenotype associations were with variants associated with blood cell trait measurements, autoimmunity and allergy, and neoplasms (**Figure 3C)**. Several mLOX specific variants are GWAS lead variants of multiple autoimmune diseases such as type 1 diabetes (rs9398805 (*CENPW*) and rs4788084 (*IL27*)), celiac disease (rs13080752 (*LPP*)), and rheumatoid arthritis (rs2371109 (*EOMES*)). Based on Open Target Genetics, none of the mLOX variants shared with mLOY were reported to be associated with any autoimmune disease. Additionally, the group of variants shared with mLOY have more associations with neoplasms (e.g., rs751343 (*ATM*) for breast cancer and rs2280548 (*MAD1L1*) for prostate cancer) and blood cell measurements than the group of variants specific for mLOX. We then examined the associations between each identified mLOX susceptibility locus and the counts of different types of blood cells^47^. Of 35 independent mLOX loci (only considering the lead variant of each locus), 33 were associated with at least one of the nine blood cell count traits examined (P<0.05), suggesting a shared genetic etiology between hematopoiesis and development of detectable mLOX (**Figure 3D**). Again, the mLOX variants shared with mLOY were among the variants associated with the most number of blood cell traits (5.5 traits average over eight variants) compared with mLOX specific variants (3 traits average over 22 variants).

To identify rare autosomal and X chromosome germline variants (MAF < 0.1%) associated with the risk of detectable mLOX, we performed gene-burden tests for our newly proposed mLOX metric which utilized information from both SNP array and WES data (mLOX 3-way combined calls) in 226,125 UKBB female participants with available WES data (**Methods**). Three non-synonymous variant functional categories were used in our analysis: high-confidence protein truncating variants (HC PTVs), missense variants with CADD scores ≥ 25 (MISS_CADD25), and damaging variants (HC_PTV+MISS_CADD25). Only one gene, *FBXO10* (F-Box Protein 10), was associated with mLOX susceptibility (P<1.2×10^−6^) (**Figure 2B**), with the strongest association observed in carriers of missense variants with CADD scores ≥25 (N of carriers=581, beta=0.059, P=1.8×10^−7^) (**Supplementary Table S17**). Logistic regression for mLOX status observed a consistent effect of *FBXO10* missense variants associated with a 2-fold increased risk of mLOX (OR=2.06 [1.59-2.68], P=1.4×10^−7^), and we further confirmed this association using a distinct analytical pipeline implementing STAAR (P=2.5×10^−7^)^48^. A leave-one-out analysis confirmed this association was not restricted to a single coding variant (P<8.5×10^−6^). *FBXO10* is the substrate-recognition component of the SCF (SKP1-CUL1-F-box protein)-type E3 ubiquitin ligase complex. The SCF (*FBXO10*) complex mediates ubiquitination and degradation of the anti-apoptotic protein, *BCL2* (BCL2 apoptosis regulator), thereby playing a role in apoptosis by controlling the stability of *BCL2*^49^.

### Allelic shift analysis for *cis* clonal selection of chromosome X alleles

As several germline variants reside on the X chromosome, we sought to investigate for a given X chromosome variant whether mLOX cells with one allele retained in a hemizygous state confers a propensity to be retained or a selective advantage over mLOX cells with the alternate X allele retained (**Figure 1B**). Conditional on mLOX having been detected, for each variant on the X chromosome, we tested whether there is a higher frequency of a given allele retained in comparison to the alternate allele being retained^14^ (**Methods**). This allelic shift analysis is similar to a transmission disequilibrium test^50^ which is robust to the presence of population structure, with only heterozygous genotypes being informative. Of the 1,645,601 X chromosome variants we examined, 25,370 (1.5%) reached the significance threshold (P<5.0×10^−8^). We identified 44 independent chromosome X variants with shifted allelic fractions on the retained X chromosome (**Methods**; **Supplementary Table S18**). The allelic shift signals spanned the length of the X chromosome (**Figure 4A**), with the strongest signals observed near the centromere (lead variant rs6612886; out of 39,246 heterozygous rs6612886 genotypes examined, 25,035 had the alternative C allele lost while 14,211 had the reference T allele lost, OR=1.76 [1.73-1.80], P=4.0×10^−659^). To investigate if the observed associations were being driven by inflation of the test statistic, we examined the relationship between the number of markers being statistically significant and the marker density within a window size of 1k bp and found no relationship between the two measures (**Supplementary Figure S10**). Similar to GWAS lead variants, 35 out of 43 lead variants (one variant dropped due to no appropriate proxy variant available in blood cell phenotype GWAS^47^) identified from allelic shift analyses were associated with at least one of blood cell phenotypes (prioritized genes *P2RY8, WAS, PJA1, PLS3, ITM2A, TMEM255A*, and *SOWAHD*), especially for several variants near the centromere region (**Figure 4B**). Finally, signals were consistent across seven biobanks further supporting the robustness of the results.

**Figure 4.**
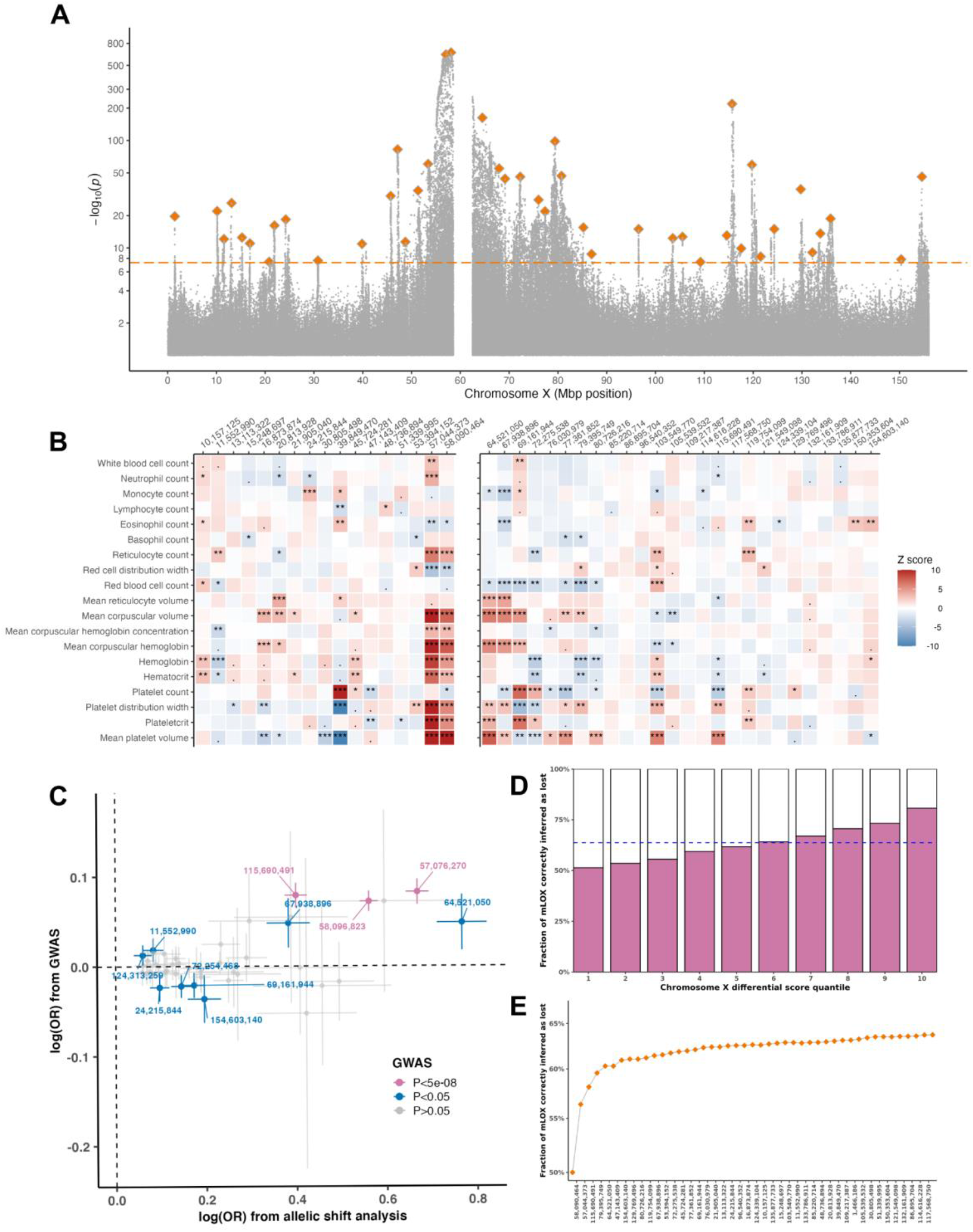
Allelic shift of chromosome X alleles among mLOX cases. Panel (**A**) shows -log_10_(P) of chromosome X variants from allelic shift analysis by meta-analyzing data of 83,320 mLOX cases from seven biobanks, with lead variants of 44 independent loci highlighted. The dashed line denotes the statistical significance (5.0×10^−8^, which is the same as the GWAS significance level) and the y axis is log scale. Panel (**B**) depicts associations of 43 allelic shift analysis lead variants with 19 blood cell phenotypes^47^. One variant was dropped due to no appropriate proxy variant available in blood cell phenotype GWAS. The absolute Z scores were cropped to the range of [0-20]. Panel (**C**) is a scatterplot of lead variants identified from allelic shift analysis (N=44) and their effects from allelic shift analysis (x axis) and GWAS (y axis). Variants are categorized based on P values from GWAS. Panel (**D**) and (**E**) show the fraction of mLOX cases with the retained X chromosome correctly inferred using an X chromosome differential score constructed from allelic shift analysis signals. To avoid overfitting, the effects of 44 lead variants were estimated from allelic shift analysis of 56,319 mLOX cases from six biobanks excluding FinnGen while the prediction performance was tested in 27,001 FinnGen mLOX cases. Panel (**D**) stratifies prediction performance by differential quantile of each X chromosome prediction score. Panel (**E**) shows the contribution of each lead variant to the prediction, starting with the most significant variants.

Among variants exhibiting significant allelic shifts in mLOX cases, 59 were missense variants (**Supplementary Table S19**) including 16 variants from 11 genes (*P2RY8, FANCB, UBA1, WAS, USP27X, VSIG4, PJA1*, CITED1, *POF1B, SAGE1*, and *MAP7D3*) likely to be lead signals (**Supplementary Figure S11**). The genes *VSIG4* (rs41307375/rs41306131 and rs17315645, r2<0.001) and *SAGE1* (rs41301507 and rs4829799, r2=0.30) each contained more than one independent missense variant. Based on the Human Protein Atlas (https://www.proteinatlas.org/), several genes with identified missense variants were also associated with cancer risk/progression (*P2RY8, UBA1, WAS*, and *SAGE1*), mental disorders (e.g., *USP27X* for intellectual disability and *PJA1* for schizophrenia^51^), or had relevance to DNA damage/repair (*FANCB*) and apoptosis (*CITED1)*. Additionally, several genes were involved in X-linked recessive disorders (e.g., *FANCB* for Fanconi anemia, *WAS* for Wiskott–Aldrich syndrome, and *POF1B* for X-linked premature ovarian failure) or known to escape from X-inactivation (e.g., *P2RY8, UBA1, WAS, VSIG4*, and *POF1B*)^3^.

Most chromosome X variants identified from the allelic shift analysis were not shared with the variants from the GWAS of mLOX (**Figure 4C**), except for rs4029980 (X:57044373:T:C, proxy SNP X:57076270:G:A, r^2^=0.87) and rs6612886 (X:58090464:T:C, proxy SNP X:58096823:A:C, r^2^=0.98) near the centromere and rs12836051 (X:115690491:A:G). Unlike GWAS, which can identify germline variants related to both chromosome missegregation and subsequent clonal selection, a large amount of chromosome X signals identified from allelic shift analysis suggests that in many women mLOX strongly favors one X chromosome over the other based on the differing allelic content of the two X chromosomes. This preference could arise from the clonal selection on retained alleles or could be due to allelic influences on X inactivation skewing (**Supplementary Figure S12**)., which later manifests as an allelic shift if mLOX occurs since mLOX affects the inactive X chromosome^10^.

We then investigated how accurately we can predict which X chromosome is likely to be retained when detectable mLOX occurs. The X chromosome differential score was constructed based on the 44 independent variants identified from allelic shift analysis by generating a chromosome-specific score for each X chromosome and calculating the difference between scores of two X chromosomes (**Methods**). To avoid overfitting, the prediction performance was tested in 27,001 FinnGen mLOX cases, with effect sizes of lead variants estimated from the allelic shift analysis of 56,319 mLOX cases from six biobanks excluding FinnGen. The fraction of mLOX cases with the retained X chromosome correctly inferred was 63.7% across all mLOX cases and up to 80.7% for mLOX cases within the top 10^th^ percentile (**Figure 4D**). When partitioning the contribution at a variant level, starting from the most significant variants (**Figure 4E**), the fraction correctly inferred reached >60% when including the first four lead variants (rs58090464, rs57044373, rs115690491, rs79395749), while the improvement of prediction accuracy from adding another 40 lead variants increased performance but was smaller in comparison (fraction from 60.3% to 63.7%). We also performed simulation analyses to assess the upper limit of prediction performance that can be reached in FinnGen mLOX cases, given the distribution of allele frequencies of 44 lead variants (**Methods**). Overall, the fraction of mLOX cases correctly inferred from real data analysis (63.7%) approached that obtained from simulation analysis (65.0%) (**Supplementary Figure S13-S14**).

## Discussion

This population-based analysis of over 900K European and Asian ancestry women indicates detectable mLOX can be observed in a substantial fraction of middle-aged and elderly women, but typically impacts less than 5% of circulating leukocytes. In an analysis of 1,253 diseases extracted from electronic health records or registry data, we identified prospective associations of mLOX with leukemia risk, specifically myeloid leukemia, and provide additional evidence for susceptibility to infectious disease. Our results indicate that the value of mLOX as a diagnostic marker could be limited to blood cancers. For non-genetic risk factors, we replicated prior mLOX associations with age and identified an association with tobacco smoking among high cell fraction mLOX. Our large sample size coupled with an improved mLOX detection approach enabled the identification of 49 common independent germline susceptibility signals across 35 loci and rare coding variations in *FBXO10* associated with mLOX. Little heterogeneity was noted in these loci across contributing studies or ancestry. The mLOX germline susceptibility signals implicate genes involved in kinetochore and spindle function, blood cell measurements, cancer predisposition, and immunity as etiologically relevant to mLOX susceptibility.

We identified shared and, more surprisingly, distinct genetic etiologies of mLOX with mLOY, which occurs frequently in aging men – albeit at higher cell fractions. The two traits are moderately correlated genome-wide and eight of the 49 mLOX variants demonstrated evidence for shared effects for both mLOX and mLOY. Shared mLOX and mLOY variants were enriched for genes important for cancer susceptibility and blood cell traits; however, effects observed for mLOX were noticeably attenuated from effects observed for mLOY. This attenuation could be due to differences in our ability to detect mLOX at lower cell fractions relative to mLOY, or could be a biological impact since mLOX is often present at lower cell fractions relative to mLOY. Variants specific to mLOX demonstrated unique evidence for associations with immunity, including HLA alleles which could play a role in the selection of X-linked cell surface antigens, as well as genes relevant to mitotic missegregation (**Supplementary Figure S15**).

In addition to conducting a GWAS, we also performed allelic shift analyses on X chromosome germline variants to identify signals of *cis* clonal selection. Allelic shift tests are similar to transmission disequilibrium tests commonly used in family trios and are robust to population stratification. These analyses identified strong independent signals of *cis* selection near the centromere as well as multiple additional signals spanning across the X chromosome. Interestingly, the majority of the allelic shift loci were not detected in the GWAS, demonstrating the ability to identify signals of selection by utilizing this approach. While the allelic shift centromeric signals were strongly associated with several blood cell phenotypes, their location near the centromere could tag germline variation with relevance for kinetochore formation and spindle attachment in this region and may predispose specific X chromosomes to missegregation errors; although, limited is known as to how germline variation in DNA sequences could impact centrosomal protein binding and spindle formation^52,53^. Other loci identified by allelic shift analyses provide support for genes involved in escaping X inactivation, cancer susceptibility, and blood cell traits as relevant to mLOX. Scores created that aggregate information across allelic shift loci correctly classified which X chromosome was more likely retained in a high percentage of mLOX women in which the difference in X chromosome scores was high. To our knowledge, this is the first demonstration of the utility of a score consisting of multiple germline variants to predict which chromosome will be lost if a somatic event occurs. Our approach for identifying variation important for chromosome X loss may be extendable to investigating other chromosomal loss events with relevance for cancer risk.

In conclusion, we provide evidence for a strong germline component to somatically occurring mLOX in which genes related to cancer susceptibility, blood cell traits, autoimmunity, and chromosomal missegregation events are relevant to mLOX susceptibility. Further, we identify many strong *cis* effects for chromosome X loci that impact which X chromosome is retained and promote clonal expansion. Genetic insights from mLOX could also be relevant to better understanding skewed X inactivation, another commonly observed X chromosome abnormality in middle-aged and elderly women.

## Supporting information

FinnGen banner authorship

BCAC funding and acknowledgment

BCAC banner authorship

MVP banner authorship

## Data Availability

Summary statistics generated from meta-analysis will be uploaded to GWAS Catalog after publication. Individual level data can be requested directly from each contributing biobank.

## Online Methods

### Definition of mosaic loss of the X chromosome (mLOX)

#### Detection of mLOX events from SNP array data in eight biobanks

All DNA samples were obtained from peripheral leukocytes and typed with single nucleotide polymorphism (SNP) arrays. The median (SD) age at sample collection for genotyping ranged from 44 (16.3) for EBB to 67.2 (12.9) for FinnGen. The calling of mosaic loss of the X chromosome (mLOX) was performed within each biobank using the Mosaic Chromosomal Alterations (MoChA) pipeline (https://github.com/freeseek/mochawdl), with GRCh38 assembly as the reference genome build. The raw genotyping array signal intensities of each variant were first transformed to B allele frequency (BAF) (relative intensity of the B allele) and Log R Ratio (LRR) (total intensity of both alleles). Then, haplotype phasing was performed using SHAPEIT4^54^ across all batches of a biobank, except for BBJ and BCAC for which phasing was done separately for each of the four sub-cohorts (cohort sizes ranged from 3,888 to 45,877 for BBJ and from 42,360 to 62,889 for BCAC). Utilizing long-range haplotype phasing can improve the sensitivity of detecting large mosaic events with low cell fractions^14^, which is characteristic of mLOX. To avoid issues with phasing and the subsequent mLOX calling, we excluded variants with poor genotyping quality such as segmental duplications with low divergence (<2%) and single-nucleotide polymorphisms (SNPs) with high levels of missingness (>3%) or heterozygote excess (P<1×10^−6^). Finally, the calling of mLOX events was performed within each batch based on the imbalance of phased BAF of heterozygous sites over the whole X chromosome. To filter out 47,XXY and 47,XXX samples, we restricted to chromosome X events with estimated ploidy less than 2.5, where the estimated ploidy is estimated by first computing the median LRR across the assayed chromosome X SNPs and then by computing the value 2^1+(LRR/LRR-hap2dip)^ with LRR-hap2dip (the difference between LRR for haploid and diploid) set at 0.45 by default. We further removed X loss events with length < 100 Mb to exclude other mosaic events (e.g., copy number neutral loss of heterozygosity) on the X chromosome. For each mLOX event that passed quality control, the fraction of cells (cf) with X loss was calculated as 4*bdev/(1+2*bdev), where bdev is the estimated BAF deviation of heterozygous sites. In addition to the dichotomous mLOX status defined by the phased BAF method, for UKBB, the mean LRR (mLRR) of variants on X chromosome non-pseudoautosomal (non-PAR) regions has also been used as a quantitative measure of mLOX.

The 2022-01-14 version of MoChA was used to detect the dichotomous mLOX status for all biobanks, except for BCAC (version: 2021-05-14) and BBJ (version: 2021-08-17 and 2021-09-07). The priors of MoChA have been updated since 2021-05-14 to improve the detection of low cell fraction mLOX calls, and thus, the biobanks that used the updated MoChA pipeline (all biobanks except for BCAC) were expected to yield higher age-adjusted mLOX prevalences than biobanks that used the previous version (only BCAC). A brief description of each contributed biobank (e.g., continental ancestry, sample size, age structures, and SNP array) is available in **Supplementary Table S1**.

#### Estimation of X chromosome dosages from UKBB whole-exome sequence data

For UKBB, the whole-exome sequence (WES) data was released in late 2021^55^, which permitted identification of X loss from sequencing allelic dosage data in combination with array data. The relative X chromosome dosage at the individual level was estimated following the steps described previously^56^. In brief, we first generated mean coverages from the original WES data for variants on the autosomes and the X chromosome non-PAR regions, separately; then, we obtained the relative X chromosome dosage by adjusting for the mean coverage of autosomes.

#### Comparison of different mLOX measures in UKBB

As mentioned above, for UKBB, three ways were used to define the mLOX phenotype, including the dichotomous mLOX status derived from the phased BAF method (by MoChA) and two quantitative measures employing either mLRR from SNP array data or allele dosage from WES data. To assess the performances of the three mLOX measures in UKBB, we compared either mLRR or X dosage between the case and the control groups defined by MoChA (**Figure S2A-C**). As shown in **Figure S2B** and **S2C**, the participants identified as mLOX cases by MoChA exhibited lower mLRR (P from the Analysis of Variance (ANOVA) test=1.5×10^−5^) and X dosage value (P<1.0×10^−250^) than mLOX controls. Then, for mLOX cases, we examined the relationships between three measures representing the extent of mosaicism (**Figure S2D-F**), including cell fraction (from MoChA), mLRR, and X dosage. Overall, significant correlations were observed across the three measures, with the absolute Pearson correlation coefficient ranging from 0.42 between mLRR and X dosage to 0.86 between mLOX cell fraction and X dosage. Again, given that mLRR is a noisier measure than X dosage, for mLOX cell fraction, a stronger correlation was observed with X dosage (r=-0.86) than with mLRR (−0.48).

### Environmental determinants and epidemiological consequences

To investigate the effect of lifestyle factors on the odds of acquiring mLOX, we assessed the associations between smoking and body mass index (BMI) with mLOX in the FinnGen cohort. In FinnGen data freeze 9, 50.3% of female participants had smoking status (N=84,926) and 18.4% had measurements for BMI (N=31,101) recorded at enrollment. We applied a logistic regression model adjusting for age (at genotyping), age^2^, and the first 10 PCs as covariates. As sensitivity analyses, we restricted the analyses to expanded mLOX calls having cf > 5%. Given that we identified a significant association between ever-smoking and expanded mLOX, we further adjusted for ever-smoking status when assessing the effect of BMI on mLOX. To examine whether the environmental determinants were shared or distinct between mLOX in women and mLOY in men, we also extended the association analyses to mLOY (N=76,808 for smoking, N=33,668 for BMI).

To assess the clinical consequences of acquiring expanded mLOX, we performed a Cox proportional hazards regression for incident cases in FinnGen, UKBB, MVP, and MGB independently, with the time-on study as the time scale. For covariates, we recommended each biobank adjust for age, age^2^, smoking, and the first 10 PCs. Meta-analysis across four biobanks was carried out with a fixed-effect model applied in the meta package^57^. For each disease, we applied Cochran’s Q-test to assess heterogeneity across biobanks with different healthcare systems. In total, we examined 1,253 phecodes covering 13 disease categories. Accordingly, the multiple-testing corrected P value threshold was set to P<4.0×10^−5^. In the main analysis, we used all detectable mLOX calls without restriction for cell fraction. For a sensitivity analysis, we considered mLOX having cf >10% as expanded calls, following the definition used by Zekavat et al^16^.

### Common and rare germline variants associated with detectable mLOX susceptibility

#### GWAS of dichotomous mLOX status in eight contributed biobanks

To identify common germline variants (minor allele frequency (MAF)>0.1%) associated with risk of detectable mLOX in peripheral leukocytes, we performed a genome-wide association study (GWAS) on chromosomes 1-22 and X in each of eight contributing biobanks independently, for a total of 904,524 women. For the dichotomous mLOX status (derived from MoChA), GWAS was conducted for FinnGen and BCAC using the Scalable and Accurate Implementation of Generalized mixed model (SAIGE)^58^ and for the other six biobanks (including UKBB) using regenie ^59^ applied in the assoc.wdl pipeline (part of the MoChA pipeline; https://github.com/freeseek/mochawdl). Both SAIGE and regenie are feasible to account for sample relatedness and extreme case-control imbalances of a dichotomous phenotype. For covariates, each biobank adjusted for age (at genotyping), age^2^, and the first 20 genetic principal components (PCs). The effective sample size, presented in Table 1, was calculated as (4*N_case_*N_control_)/(N_case_+N_control_).

#### GWAS of 3-way combined quantitative mLOX measure in UKBB

For UKBB, to improve the power of GWAS, we proposed a new quantitative measure by combining the three methods of mLOX calling, that is, the mLOX combined call (3-way) = mLOX-status + 2*cf - 2*mLRR - 4*(dosage-2) (cropped to the range [0,2]). The intuition behind this newly proposed measure was to emphasize mLOX cases with larger cell fractions (similar to the strategy used by a recent mosaic loss of the Y chromosome (mLOY) study^60^) while obtaining enhanced mLOX calls from integrating independent information of both SNP array and WES data. Compared to the dichotomous mLOX status derived from MoChA, the t-test statistic for association with age was increased by 29.2% when using the 3-way combined calls. As not all participants with SNP array data had WES data available, we imputed the missing 3-way mLOX combined calls with 2-way combined calls, defined as mLOX-status + 3*cf– 3*mLRR (cropped to the range [0,2] as well). For the proposed quantitative mLOX measure, GWAS was performed with the linear mixed model applied in BOLT-LMM^61^.

#### Meta-analysis

For each contributed biobank, we filtered out variants with MAF < 0.1% or imputation INFO score < 0.6. We also inspected allele frequencies of each biobank versus Genome Aggregation Database (gnomAD) 3.0 as well as the relationship between standard errors and effective sample sizes across biobanks, as applied by the covid-19 HGI meta-analysis^62^. Given that no biobank deviated from the expected pattern, we conducted meta-analyses across biobanks. In addition to the dichotomous mLOX measure used by all biobanks, UKBB was able to run GWAS with an additional quantitative measure that combined information of three ways of mLOX calling and thus was expected to yield increased power in GWAS. Depending on which mLOX measure was used in the UKBB GWAS, we applied two fixed-effect meta-analysis models accordingly. When using the dichotomous measure, we applied the inverse variance weighting (IVW) method which weighted the effect size estimated from an individual biobank by its inverse variance. When UKBB used the 3-way combined measure as the GWAS phenotype, we employed the weighted z-score method (weighted by the square root of the effective sample size) applied in the METAL software^63^ which can manage the different units of dichotomous and quantitative measures. As the main analysis, we meta-analyzed summary statistics across all eight biobanks regardless of ancestry and applied Cochran’s Q-test to assess the heterogeneity. To further investigate the impact of ancestry, we also conducted a meta-analysis for 7 biobanks containing only participants of European ancestry (without BBJ of East Asian ancestry).

#### Independent loci identification and gene prioritization

To identify independent signals and prioritize candidate causal genes, we applied the GWAStoGenes pipeline for variants presented in at least half of the contributed biobanks. In brief, primary independent signals associated with mLOX susceptibility at a genome-wide significant level (P<5×10^−8^) were initially selected in 1Mb windows. Secondary independent signals were identified by using an approximate conditional analysis applied in GCTA^64^, with LD structures constructed from UKBB samples. Secondary signals were only considered if they were genome-wide significant, in low LD (r2<0.05) with primary signals, and having association statistics unchanged with the conditional analysis. We also excluded variants without any nearby genes (within 500 kb) documented in the NCBI RefSeq dataset^65^.

Candidate genes were prioritized using the following criteria and scored by their strength of evidence for causality. First, signals were annotated with their physically closest genes. Second, signals and their closely linked variants (R^2^ > 0.8) were annotated if they were predicted deleterious coding variants, or if the paired genes exhibited a gene-level association when collapsing all predicted deleterious coding variants within a gene using Multi-marker Analysis of GenoMic Annotation (MAGMA)^66^. Third, non-coding signals and closely-linked variants were then annotated if they could be mapped to known enhancers via the activity-by-contact (ABC) model^67^. Fourth, colocalization between GWAS and expression quantitative trait locus (eQTL) data was performed using the summary data-based Mendelian randomization (SMR) and heterogeneity in dependent instruments (HEIDI) test (version 0.68)^68^ and the Approximate Bayes Factor (ABF) method applied in the “coloc” package (version 5.1.0)^69^. To identify tissues exhibiting a significant genome-wide enrichment, we used LD score regression applied to specifically expressed gene (LDSC-SEG)^70^ approach, with eQTL datasets from cross-tissue meta-analyzed GTEx eQTL v.7^71^, eQTLGen^72^, and Brain-eMeta^73^. The same set of analyses were also applied to a protein quantitative trait locus (pQTL) dataset^74^. Finally, by integrating GWAS summary statistics with data from gene expression, biological pathway, and predicted protein-protein interaction, candidate genes were identified using the gene-level Polygenic Priority Score (PoPS) method^75^.

#### Gene-burden test for rare variants causing detectable mLOX

To identify rare germline variants (minor allele frequency (MAF) < 0.1%) associated with the risk of detectable mLOX, we performed gene-burden tests on chromosomes 1-22 and X in 226,125 UKBB female participants with WES data available. We performed WES data pre-processing and quality control following Gardner et al.^76^. We annotated variants using the ENSEMBL Variant Effect Predictor (VEP) v104^77^ and defined protein-truncating variants (PTVs) as high-confidence (HC, as defined by LOFTEE) stop gained, splice donor/acceptor, and frameshift consequences. We then utilized CADDv1.6 to score a variant based on its predicted deleteriousness^78^. Only non-synonymous variants with MAF < 0.1% were included in the analysis. As the main analysis, we used BOLT-LMM^61^ to perform the gene-burden test. For each gene, we defined individuals with HC PTVs, missense variants with CADD scores ≥ 25 (MISS_CADD25), and damaging variants (HC_PTV + MISS_CADD25) (DMG) as carriers. Then, carriers with non-synonymous variants were defined as heterozygous and non-carriers as homozygous. For covariates, we adjusted for age, age^2^, batches, sex, and the first ten PCs. We further excluded the genes with less than 50 non-synonymous variant carriers for each setting, resulting in 8,702 genes for HC_PTV, 15,144 for MISS_CADD25, and 16,493 for DMG, for a total of 40,339 genes. Accordingly, the Bonferroni corrected exome-wide significant threshold was set to 0.05/40,339=1.24×10^−6^. To avoid the identified association dominated by a single variant, as sensitivity analysis, we conducted a leave-one-out analysis using a generalized linear model for each significant gene. In addition, we reproduced the associations detected by BOLT-LMM with STAAR^48^.

#### Pathway and gene set analysis

To identify gene sets enriched in the same biological process, we performed pathway-based analysis using the summary data-based adaptive rank truncated product (sARTP) method^79^. We used summary statistics from meta-analysis of seven biobanks of European ancestry (without BBJ) and LD structures constructed from European ancestry samples of the 1000 Genomes project (1000 Genomes Project Consortium, Nature, 2015). We considered a total of 6,285 gene sets available in GSEA (https://www.gseamsigdb.org/gsea/msigdb/). Accordingly, the Bonferroni corrected P value was set to 0.05/6,285=8.0×10^−6^.

#### Genetic correlation

To investigate whether there are traits that are genetically correlated with mLOX susceptibility, we estimated genetic correlations between mLOX and 60 phenotypes (including both major diseases and blood cell phenotypes) using LD score regression (LDSC)^80^. For LDSC, we used HapMap3^81^ SNPs and LD structures constructed from 1000 Genomes project^82^ samples of European ancestry.

#### Per-chromosome heritability

To examine whether the observed heritability for each chromosome was proportional to chromosome length, we estimated per-chromosome heritability for 3-way combined mLOX measure in UKBB using BOLT-REML^83^. Given the large associations of HLA genes, we further examined how heritability explained by chromosome 6 changed after excluding variants from the extended MHC region (GRCh38: chr6:25.7-33.4 Mb).

### Shared and distinct mechanisms between mLOX in women and mLOY in men

#### Bayesian models to cluster variants by effects on mLOX and mLOY

We employed a Bayesian line model framework (https://github.com/mjpirinen/linemodels) to assign each of the 49 independent common variants identified from mLOX GWAS and 147 variants (nine variants dropped due to missing in mLOX GWAS) from the published mLOY GWAS^13^ into three groups: specific to mLOX, specific to mLOY, and shared between mLOX and mLOY. The slopes of the line models were set to 0 for the group of variants specific for mLOY and infinite for variants specific for mLOX. For variants shared between mLOX and mLOY, the slope was set to 0.3, based on the effects of four variants (rs568868093, rs381500, rs2280548, rs78378222) that were genome-wide significant in both mLOX GWAS and mLOY GWAS. For all three line models, the prior SD determining the magnitude of the effects was set to 0.15 and the correlation parameter determining the allowed deviations from the lines to 0.995. The correlation between mLOX and mLOY GWAS statistics was set to 0 given that there was no overlap between samples used in the two GWAS. We assumed a uniform prior for the three models and obtained the posterior probabilities for each data point separately within a Bayesian framework. Probability assignment threshold was set to 95%.

#### Fine-mapping of HLA alleles for mLOX and mLOY in FinnGen

Given the large associations with mLOX and the high polymorphism of HLA genes, we fine-mapped HLA alleles at a unique protein sequence level in the FinnGen cohort. In FinnGen data freeze 9, a total of 172 HLA alleles of 10 transplantation genes were imputed using a Finnish-specific reference panel, as described in Ritari et al.^84^. We conducted the association analysis between each imputed HLA allele and the dichotomous mLOX status in 168,838 Finnish female participants (N of cases = 27,001) using a multivariate logistic regression model, considering age, age^2^, and the first 10 PCs as covariates. Only HLA alleles with more than 5 mLOX cases carrying the minor alleles were included in the analysis. Ultimately, we considered 156 HLA alleles for mLOX, including 18 alleles for HLA-A, 36 for HLA-B, 20 for HLA-C, 29 for HLA-DRB1, 14 for HLA-DQA1, 14 for HLA-DQB1, 18 for HLA-DPB1, 3 for HLA-DRB3, and 2 each for HLA-DRB4, and DRB5. To identify independent HLA alleles, a stepwise conditional analysis was performed with each step adding the most significant HLA allele obtained from the previous step as an additional covariate, until no HLA allele can reach the significant threshold. To examine whether the HLA associations are shared between mLOX and mLOY, we extended the HLA fine-mapping analyses to mLOY in men (total N = 128,729, N of cases = 45,675) for 157 HLA alleles (including HLA-A*02:02 compared to the 156 alleles used by mLOX association analyses).

### Allelic shift analysis for *cis* clonal selection of chromosome X alleles

#### Allelic shift analysis

Conditional on mLOX having been detected, for each variant on the X chromosome we tested whether there is a propensity for X chromosomes with a given allele to be identified as lost more often than X chromosomes with the other allele. Similar to a transmission disequilibrium test^50^, this test is robust to the presence of population structure. Rather than measuring the over-transmission of an allele from heterozygous parents to offspring, we measured the propensity of alleles to be on the retained chromosome X homologue. Therefore, we carried out a binomial test for each variant with a sample size equal to the number of women with detected mLOX who were heterozygous for that variant, with no need to correct for covariates or relatedness.

Given the large number of X chromosome signals observed from the allelic shift analysis, we inspected whether inflation may have contributed to the signals. We hypothesized that if the signals were random, then the number of variants being significant can be related to the number of variants in that region. Therefore, we checked the number of variants per 1kb region across the whole X chromosome.

#### Identification of independent loci

Given the complexity of LD structures for X chromosomes especially for centromere and pseudoautosomal (PAR) regions, we defined index variants by iteratively spanning the ± 500 kb region around the most significant variant until no further variants reached a genome-wide significant level (P<5×10^−8^). Then, we calculated LD between every two index variants and kept the variant with a lower P value if a pair of index variants with r2<0.1.

#### Polygenic score to predict the retained X chromosome

To assess how well the allelic shift analysis polygenic score (PGS) can predict which X chromosome is retained when mLOX occurs, we constructed PGSs in FinnGen mLOX cases (N=27,001). In brief, we extracted the effect size for 44 independent loci from the allelic shift analysis of 6 biobanks excluding FinnGen. Given that MoChA was able to detect which alleles were lost at heterozygous sites, for each mLOX case, we computed the PGS for the retained X chromosome (PGS_retained_) and the lost X chromosome (PGS_lost_) separately and obtained the difference in PGS between the two X chromosomes (PGS_diff_=PGS_lost_-PGS_retained_). A negative PGS_diff_ indicates that the retained X chromosome of the mLOX case was correctly predicted. To assess the upper limit of prediction performance, we performed simulation analysis in FinnGen mLOX cases based on the distribution of allele frequencies of 44 lead variants in the Finnish population.

## Code availability

The Mosaic Chromosomal Alterations (MoChA) pipelines used for mosaic loss of the X chromosome calling (mocha.wdl), GWAS (assoc.wdl), allelic shift analysis (impute.wdl and shift.wdl), and X chromosome differential score estimation (score.wdl) are available at https://github.com/freeseek/mochawdl. The GWAS meta-analysis was performed by using the pipeline developed by COVID-19 HGI, available at https://github.com/covid19-hg/META_ANALYSIS.

## Acknowledgments

We thank Juha Karjalainen (Institute for Molecular Medicine Finland (FIMM), Finland) and Mattia Cordioli (FIMM, Finland) for assistance in GWAS meta-analysis, Shea J. Andrews (Icahn School of Medicine at Mount Sinai, USA) and Jaakko Leinonen (FIMM, Finland) for kindly sharing formatted GWAS summary statistics used in genetic correlation analyses, Sakari Jukarainen (FIMM, Finland) and Alessio Gerussi (University of Milano-Bicocca, Italy) for insightful discussion on pheWAS analyses from a clinical standpoint, Samuel Jones (FIMM, Finland) and Masahiro Kanai (Broad Institute of MIT and Harvard, USA) for valuable feedback on HLA and fine-mapping, Jukka Koskela (FIMM, Finland) and Mikko Myllymäki (FIMM, Finland) for discussion on clonal hematopoiesis, Yu Fu (FIMM, Finland) and Annina Preussner (FIMM, Finland) for discussion on genetic analyses of sex chromosomes, and Geert Kops for discussion on mechanism causing chromosome missegregation. We thank Ms. Azusa Kouno in RIKEN Center for Integrative Medical Sciences and the members of the BioBank Japan Project, headquartered in the University of Tokyo Institute of Medical Science, for supporting this project. We want to acknowledge the participants and investigators of each contributing biobank. The FinnGen project is funded by two grants from Business Finland (HUS 4685/31/2016 and UH 4386/31/2016) and the following industry partners: AbbVie Inc., AstraZeneca UK Ltd, Biogen MA Inc., Bristol Myers Squibb (and Celgene Corporation & Celgene International II Sàrl), Genentech Inc., Merck Sharp & Dohme LCC, Pfizer Inc., GlaxoSmithKline Intellectual Property Development Ltd., Sanofi US Services Inc., Maze Therapeutics Inc., Janssen Biotech Inc, Novartis AG, and Boehringer Ingelheim International GmbH. Following biobanks are acknowledged for delivering biobank samples to FinnGen: Auria Biobank (www.auria.fi/biopankki), THL Biobank (www.thl.fi/biobank), Helsinki Biobank (www.helsinginbiopankki.fi), Biobank Borealis of Northern Finland (https://www.ppshp.fi/Tutkimus-ja-opetus/Biopankki/Pages/Biobank-Borealis-briefly-in-English.aspx), Finnish Clinical Biobank Tampere (www.tays.fi/en-US/Research_and_development/Finnish_Clinical_Biobank_Tampere), Biobank of Eastern Finland (www.ita-suomenbiopankki.fi/en), Central Finland Biobank (www.ksshp.fi/fi-FI/Potilaalle/Biopankki), Finnish Red Cross Blood Service Biobank (www.veripalvelu.fi/verenluovutus/biopankkitoiminta), Terveystalo Biobank (www.terveystalo.com/fi/Yritystietoa/Terveystalo-Biopankki/Biopankki/) and Arctic Biobank (https://www.oulu.fi/en/university/faculties-and-units/faculty-medicine/northern-finland-birth-cohorts-and-arctic-biobank). All Finnish Biobanks are members of BBMRI.fi infrastructure (www.bbmri.fi). Finnish Biobank Cooperative -FINBB (https://finbb.fi/) is the coordinator of BBMRI-ERIC operations in Finland. The Finnish biobank data can be accessed through the Fingenious® services (https://site.fingenious.fi/en/) managed by FINBB. For BCAC and MVP, the detailed acknowledgement is available in Supplementary materials.

## Author contributions

This project is initialized and led by A.L., G.G., P.-R.L, A.G., J.R.B.P., and M.M.. A.L. and M.M. wrote the first draft of the manuscript. A.L. coordinated the analyses of each contributing biobank, performed FinnGen specific analyses, conducted meta-analysis (including GWAS, allelic shift analysis, and pheWAS) and post-GWAS analyses, generated the figures and tables, and wrote the manuscript. G.G. developed the MoChA pipelines for mLOX calling, GWAS, allelic shift analysis, and X chromosome differential score estimation, guided the analyses of each contributing biobank, performed mLOX calling, GWAS, and allelic shift analysis for UKBB and MGB, and wrote the manuscript. Y.Z. performed WES analyses and 3-way combined call GWAS in UKBB, generated Supplementary Figure S2 and S6, prepared Supplementary Table S17, and wrote the relevant result and method paragraphs. M.P developed the Bayesian line model to cluster mLOX and mLOY loci and wrote the relevant method paragraph. M.M.Z. performed pheWAS for UKBB, MGB, and MVP and GWAS for MGB. K.K. performed the GWAS to gene pipeline, prepared Supplementary Table S11, and wrote the relevant method paragraphs. Z.Y. estimated heritability and genetic correlations and prepared Supplementary Table S14. K.Y. and L.S. performed the pathway analysis and prepared Supplementary Table S12. C.V. performed the sensitivity analyses for associations with leukemia in UKBB and prepared Supplementary Table S7. X.L. performed mLOX calling, GWAS, allelic shift analysis, and HLA fine-mapping replication analysis in BBJ. D.W.B. performed GWAS for PLCO and generated inputs for blood cell trait heat-map (Figure 3D and 4B).

G.H. performed mLOX calling, GWAS, and allelic shift analysis for EBB. B.G. and S.P. performed mLOX calling, GWAS, and allelic shift analysis for MVP. J.D performed mLOX calling and GWAS for BCAC. W.Z. performed mLOX calling, GWAS, and allelic shift analysis for PLCO. Y.M. participated in BBJ analyses. V.T. and F.-D.P participated in EBB analyses. M.A., T.P.S, and A.G. participated in FinnGen analyses. W-Y.H. and N.F. participated in PLCO analyses. E.J.G. participated in UKBB WES analyses. V.G.S. assisted in interpretating findings related to clonal hematopoiesis. A.P. coordinated the FinnGen project. H.M.O advised the HLA fine-mapping analysis and assisted in interpretating findings related to HLA. T.T. assisted in interpretating findings related to skewed X-chromosome inactivation and escaping from X-chromosome inactivation. S.J.C. coordinated the PLCO project. R.M. supervised EBB analyses. P.N. supervised pheWAS for UKBB, MGB, and MVP. M.J.D. initialized/conceptualized the mosaic chromosomal alteration project in FinnGen and assisted in interpretating findings especially those related to mLOY in men. A.B. supervised pheWAS in UKBB, MGB, and MVP and the sensitivity analyses for associations with leukemia in UKBB. S.A.M. supervised the development of MoChA pipelines. C.T. supervised BBJ analyses and advised the HLA fine-mapping analysis. P.-R.L., A.G., J.R.B.P, and M.M. co-supervised the project, interpreted the findings, and wrote the manuscript. For FinnGen, BCAC, and MVP, detailed author lists are available in supplementary materials. All authors reviewed the manuscript.

## Ethics statement

Patients and control subjects in FinnGen provided informed consent for biobank research, based on the Finnish Biobank Act. Alternatively, separate research cohorts, collected prior the Finnish Biobank Act came into effect (in September 2013) and start of FinnGen (August 2017), were collected based on study-specific consents and later transferred to the Finnish biobanks after approval by Fimea (Finnish Medicines Agency), the National Supervisory Authority for Welfare and Health. Recruitment protocols followed the biobank protocols approved by Fimea. The Coordinating Ethics Committee of the Hospital District of Helsinki and Uusimaa (HUS) statement number for the FinnGen study is Nr HUS/990/2017. The FinnGen study is approved by Finnish Institute for Health and Welfare (permit numbers: THL/2031/6.02.00/2017, THL/1101/5.05.00/2017, THL/341/6.02.00/2018, THL/2222/6.02.00/2018, THL/283/6.02.00/2019, THL/1721/5.05.00/2019 and THL/1524/5.05.00/2020), Digital and population data service agency (permit numbers: VRK43431/2017-3, VRK/6909/2018-3, VRK/4415/2019-3), the Social Insurance Institution (permit numbers: KELA 58/522/2017, KELA 131/522/2018, KELA 70/522/2019, KELA 98/522/2019, KELA 134/522/2019, KELA 138/522/2019, KELA 2/522/2020, KELA 16/522/2020), Findata permit numbers THL/2364/14.02/2020, THL/4055/14.06.00/2020,,THL/3433/14.06.00/2020, THL/4432/14.06/2020, THL/5189/14.06/2020, THL/5894/14.06.00/2020, THL/6619/14.06.00/2020, THL/209/14.06.00/2021, THL/688/14.06.00/2021, THL/1284/14.06.00/2021, THL/1965/14.06.00/2021, THL/5546/14.02.00/2020, THL/2658/14.06.00/2021, THL/4235/14.06.00/202, Statistics Finland (permit numbers: TK-53-1041-17 and TK/143/07.03.00/2020 (earlier TK-53-90-20) TK/1735/07.03.00/2021, TK/3112/07.03.00/2021) and Finnish Registry for Kidney Diseases permission/extract from the meeting minutes on 4^th^ July 2019. The Biobank Access Decisions for FinnGen samples and data utilized in FinnGen Data Freeze 9 include: THL Biobank BB2017_55, BB2017_111, BB2018_19, BB_2018_34, BB_2018_67, BB2018_71, BB2019_7, BB2019_8, BB2019_26, BB2020_1, Finnish Red Cross Blood Service Biobank 7.12.2017, Helsinki Biobank HUS/359/2017, HUS/248/2020, Auria Biobank AB17-5154 and amendment #1 (August 17 2020), AB20-5926 and amendment #1 (April 23 2020) and it’s modification (Sep 22 2021), Biobank Borealis of Northern Finland_2017_1013, Biobank of Eastern Finland 1186/2018 and amendment 22 § /2020, Finnish Clinical Biobank Tampere MH0004 and amendments (21.02.2020 & 06.10.2020), Central Finland Biobank 1-2017, and Terveystalo Biobank STB 2018001 and amendment 25^th^ Aug 2020. The UKBB analyses were conducted using applications 7089, 9905, and 21552.

