## Supplemental Figure S1-S15 for "Population analyses of mosaic X chromosome loss identify genetic drivers and widespread signatures of cellular selection"

^1^Institute for Molecular Medicine Finland (FIMM), HiLIFE, University of Helsinki, Helsinki, Finland. ^2^Program in Medical and Population Genetics, Broad Institute of MIT and Harvard, Cambridge, MA, USA. ^3^Stanley Center, Broad Institute of Harvard and MIT, Cambridge, MA, USA. ^4^Department of Genetics, Harvard Medical School, Boston, MA, USA. ^5^MRC Epidemiology Unit, Institute of Metabolic Science, University of Cambridge, Cambridge, UK. ^6^Department of Publich Health, University of Helsinki, Helsinki, Finland. ^7^Department of Mathematics and Statistics, University of Helsinki, Helsinki, Finland. ^8^Cardiovascular Research Center, Massachusetts General Hospital, Boston, MA, USA. ^9^Division of Cancer Epidemiology and Genetics, National Cancer Institute, Rockville, MD, USA. ^10^Department of Medicine, Queen’s University, Kingston, ON, Canada. ^11^Laboratory for Statistical and Translational Genetics, RIKEN Center for Integrative Medical Sciences, Yokohama, Japan. ^12^Cancer Prevention Fellowship Program, Division of Cancer Prevention, National Cancer Institute, Rockville, MD, USA. ^13^Estonian Genome Centre, Institute of Genomics, University of Tartu, Tartu, Estonia. ^14^Center for Data and Computational Sciences (C-DACS), VA Cooperative Studies Program, VA Boston Healthcare System, Boston, MA, USA. ^15^Booz Allen Hamilton, McLean, VA, USA. ^16^Centre for Cancer Genetic Epidemiology, Department of Public Health and Primary Care, University of Cambridge, Cambridge, UK. ^17^Laboratory for Genotyping Development, RIKEN Center for Integrative Medical Sciences, Yokohama, Japan. ^18^Department of Medicine, Brigham and Women’s Hospital, Harvard Medical School, Boston, MA, USA. ^19^Division of Hematology/Oncology, Boston Children’s Hospital, Harvard Medical School, Boston, MA, USA. ^20^Department of Pediatric Oncology, Dana-Farber Cancer Institute, Harvard Medical School, Boston, MA, USA. ^21^Analytic and Translational Genetics Unit, Massachusetts General Hospital, Boston, MA, USA. ^22^Anesthesia, Critical Care, and Pain Medicine, Massachusetts General Hospital, Boston, MA, USA. ^23^Center of Genomic Medicine, Massachusetts General Hospital, Boston, MA, USA. ^24^Division of Genetic Medicine, Department of Medicine, Vanderbilt University Medical Center, Nashville, TN, USA. ^25^Clinical Research Center, Shizuoka General Hospital, Shizuoka, Japan. ^26^Department of Applied Genetics, School of Pharmaceutical Sciences, University of Shizuoka, Shizuoka, Japan. ^27^Center for Data Sciences, Brigham and Women’s Hospital, Harvard Medical School, Boston, MA, USA. ^28^These authors contributed equally: Aoxing Liu, Giulio Genovese, Yajie Zhao. ^29^These authors jointly supervised this work: Po-Ru Loh, Andrea Ganna, John R.B. Perry, Mitchell J. Machiela.

**Figure S1. Cell fraction of detected mosaic loss of the X chromosome (mLOX) in females and mosaic loss of the Y chromosome (mLOY) in males of FinnGen.**

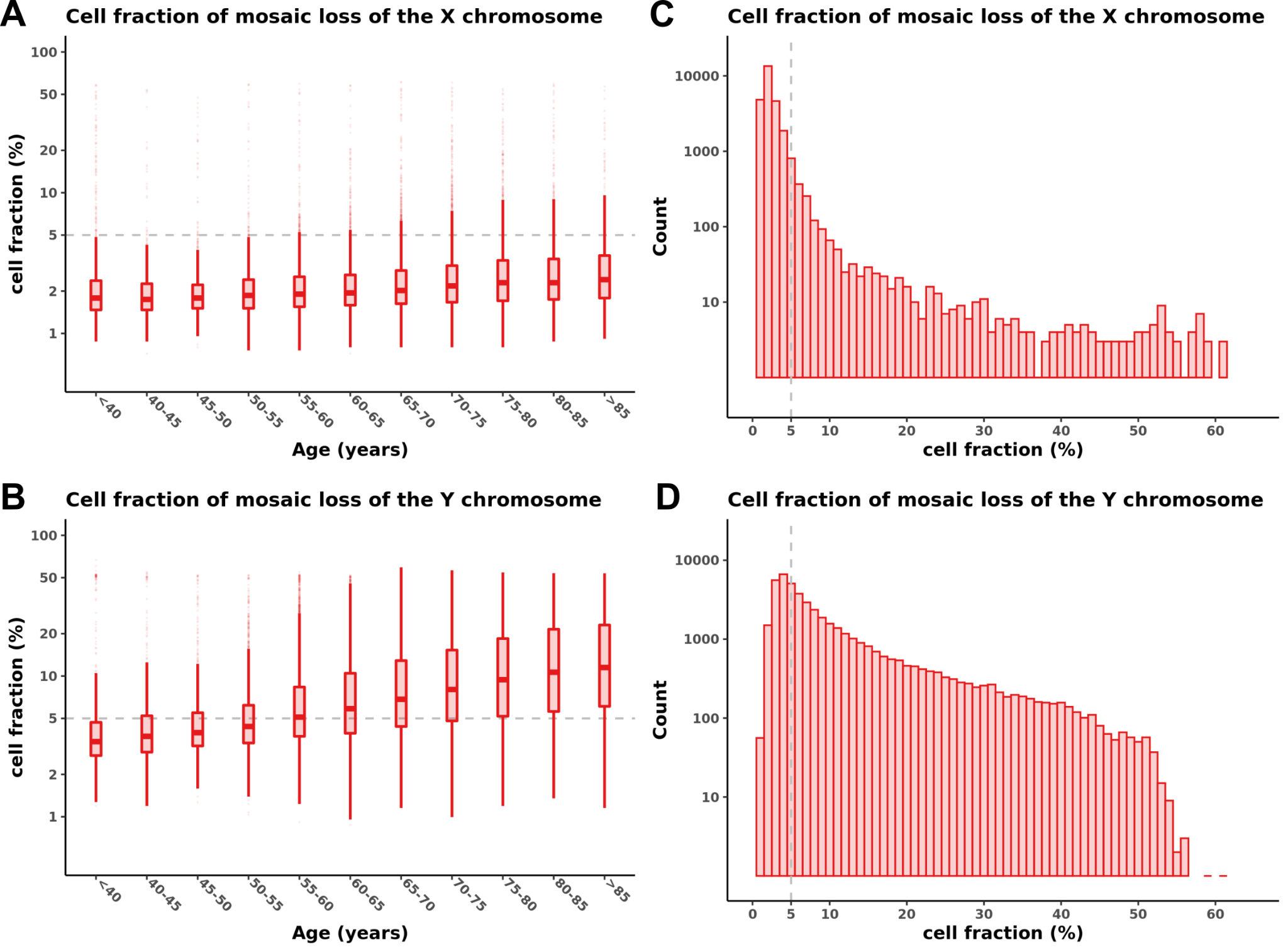

Panel (**A**) and (**B**) show the distribution of cell fractions of mosaic loss events by age at DNA sample collection in 27,001 FinnGen mLOX cases and 45,670 mLOY cases. Panel (**C**) and (**D**) show counts of mLOX or mLOY samples by cell fraction. The dashed line, corresponding to 5% cell fraction, represents the threshold for expanded calls. Pearson correlation coefficient between age at DNA sample collection and cell fraction was 5.0% [3.8%-6.2%] (P=3.3e-16) in mLOX cases but reached 25.8% [24.9%, 26.6%] (P<2.2e-16) in mLOY cases. The lower correlation between mLOX cell fraction and genotyping age could be due to that, mLOX cases have an overall much lower cell fraction (median=2.0 % for mLOX and 6.5% for mLOY) and limited variations in cell fraction (SD=3.7% for mLOX and 9.6% for mLOY) compared to mLOY cases.

**Figure S2. Estimates of X chromosome intensity (mLRR-X) from SNP array data and dosage from whole-exome sequence (WES) data in UKBB female participants, stratified by the dichotomous mosaic loss of the X chromosome (mLOX) status classified by the MoChA pipeline.**

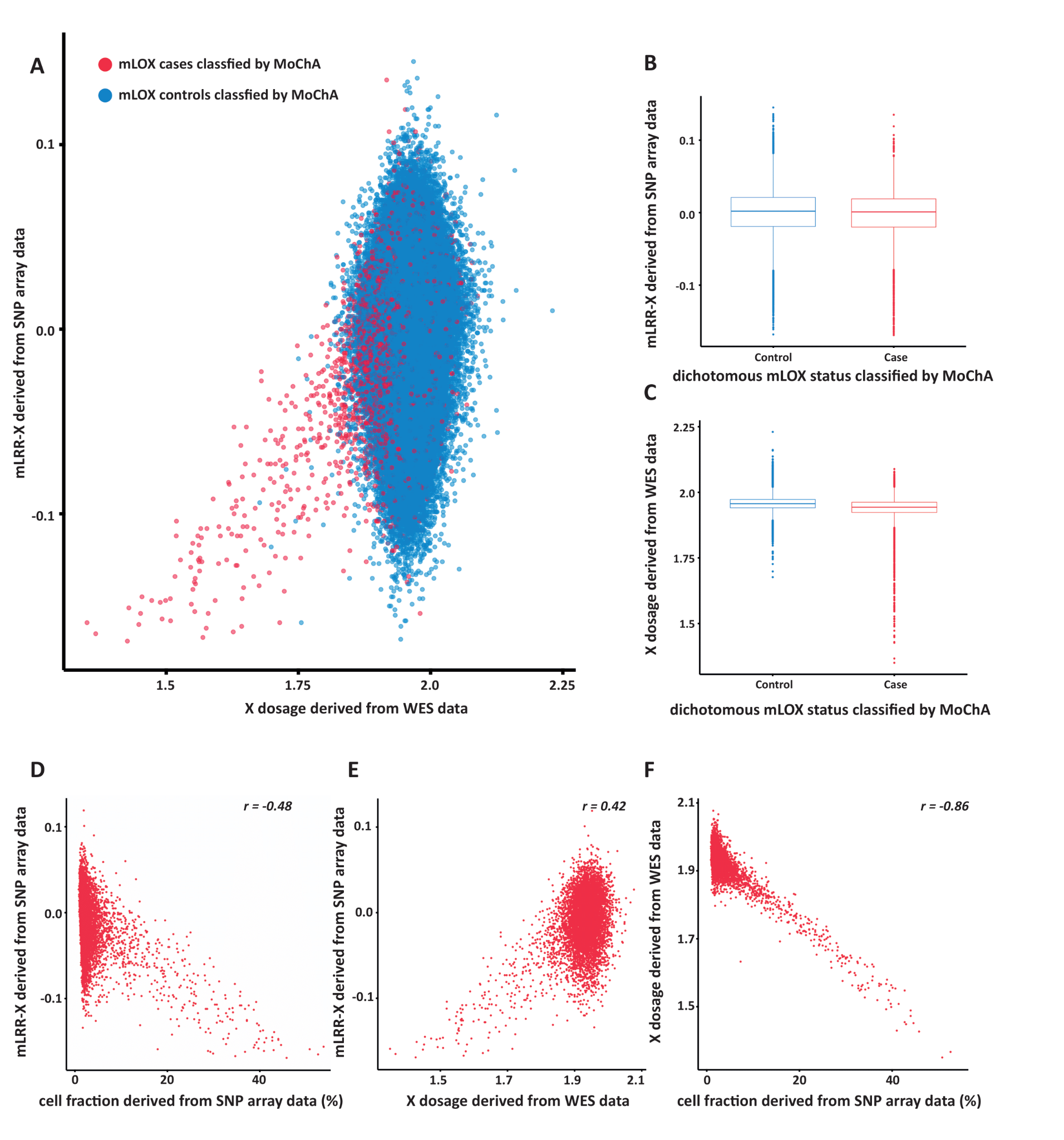

Panel (**A**) shows the relationship between X chromosome dosage and mLRR-X in UKBB female participants with both SNP array and WES data available (N=243,520), stratified by the dichotomous mLOX status classified by the MoChA pipeline. Panel (**B**) and (**C**) compare either mLRR-X or X dosage between the mLOX case and the control groups. Panel (**D**), (**E**), (**F**) show the relationships between cell fraction (estimated BAF deviation of heterozygous sites from SNP array data), mLRR-X, and X dosage in mLOX cases.

**Figure S3. Prevalence of mosaic loss of the X chromosome (mLOX) by age at genotyping in each contributed biobank.**

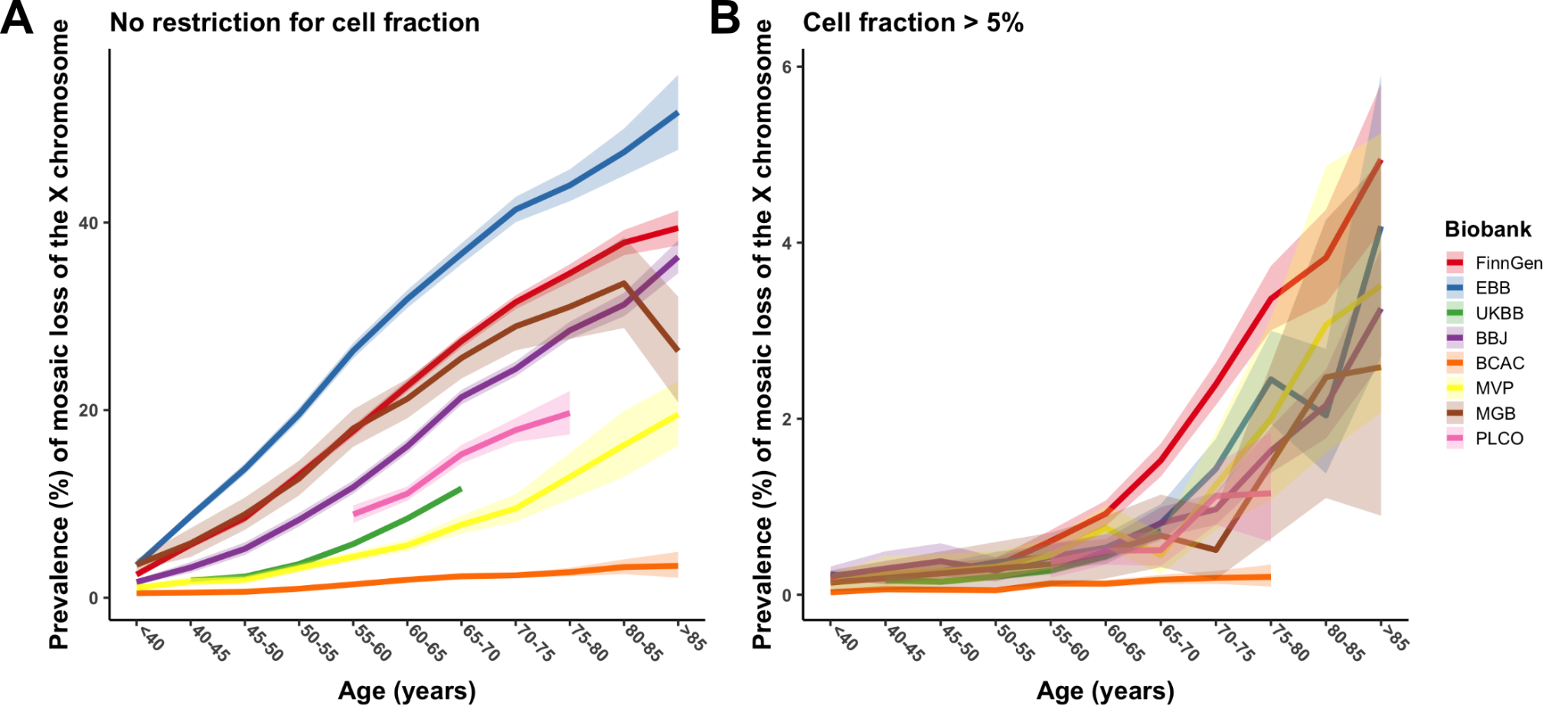

**Figure S4. Prevalence of detectable mosaic loss of the X chromosome (mLOX) by age at genotyping in females from FinnGen, stratified by ever-smoking status.**

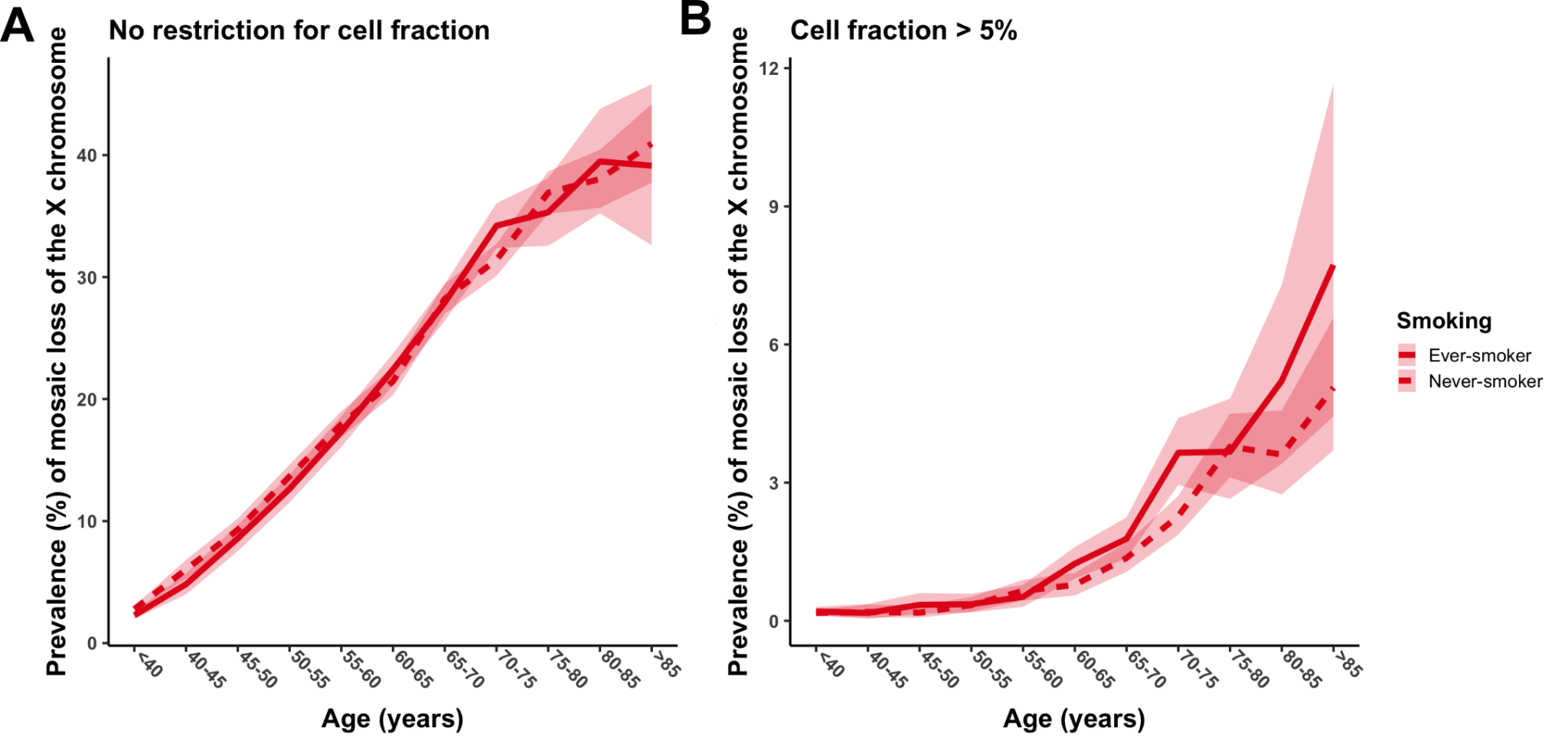

**Figure S5. Associations of ever-smoking behavior with detectable mosaic loss of the X chromosome (mLOX) in females from FinnGen, stratified by prevalent cancer status.**

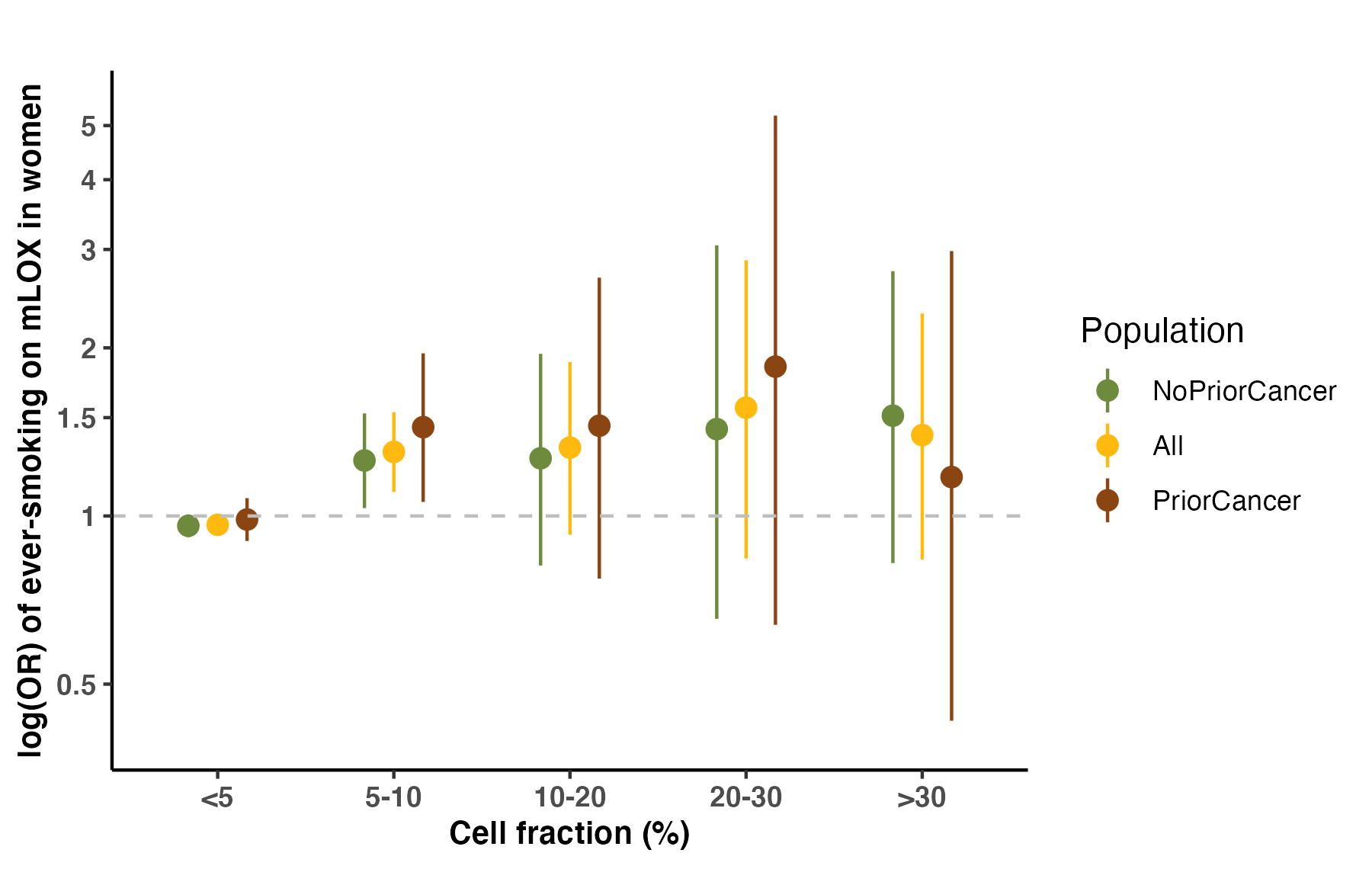

**Figure S6. Per-chromosome heritability for 3-way combined mosaic loss of the X chromosome (mLOX) measure in UKBB.**

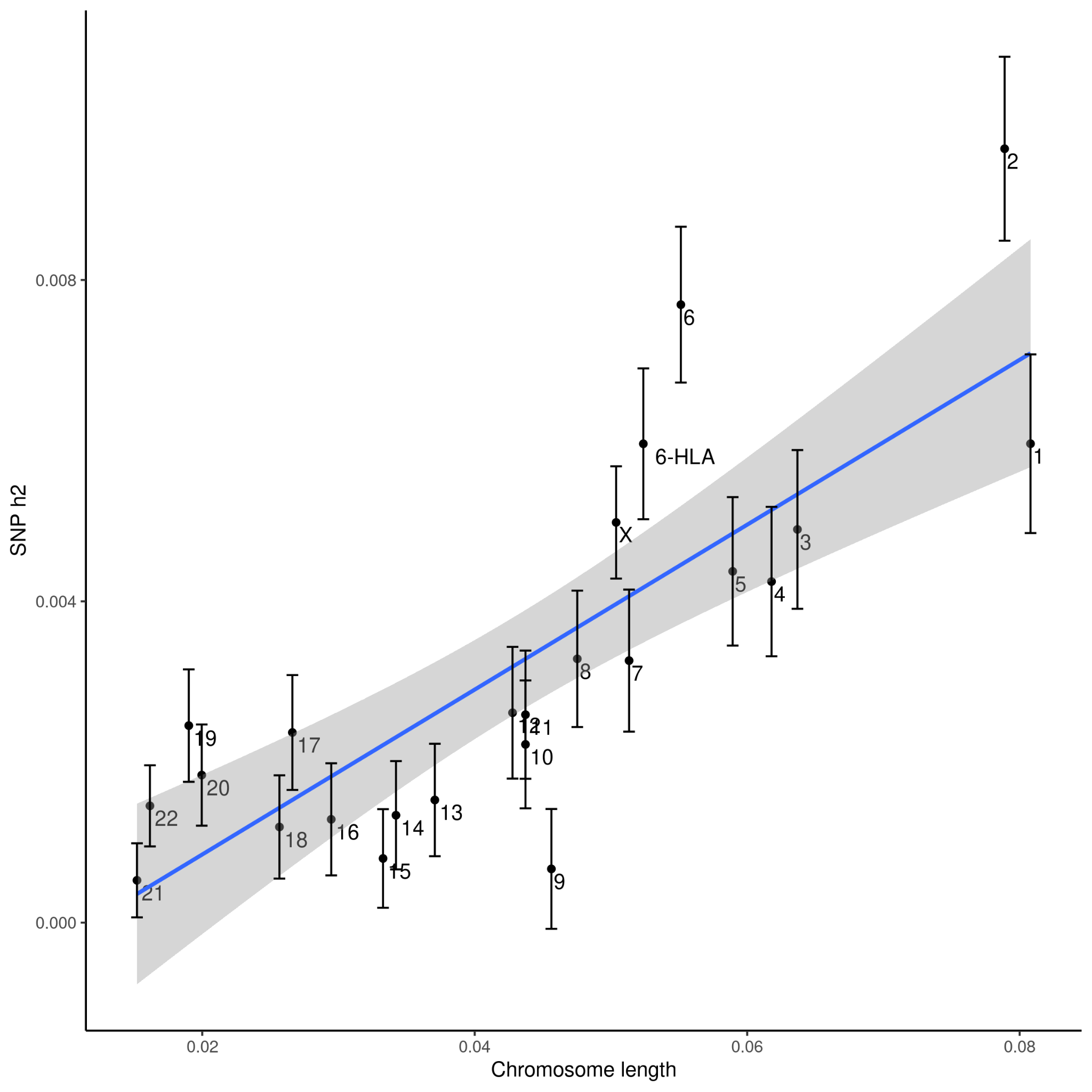

When partitioning heritability by chromosome in UKBB, we observed that, chromosome 2, 6, and X explained more heritability than expected by its chromosome length. For chromosome 6, the explained heritability dropped from 0.0077 (0.001) to 0.006 (0.0009) after excluding variants from the extended MHC region (GRCh38: chr6:25.7-33.4 Mb).

**Figure S7. Effects of mosaic loss of the X chromosome (mLOX) susceptibility for lead variants identified from GWAS meta-analysis, stratified by cell fractions in FinnGen.**

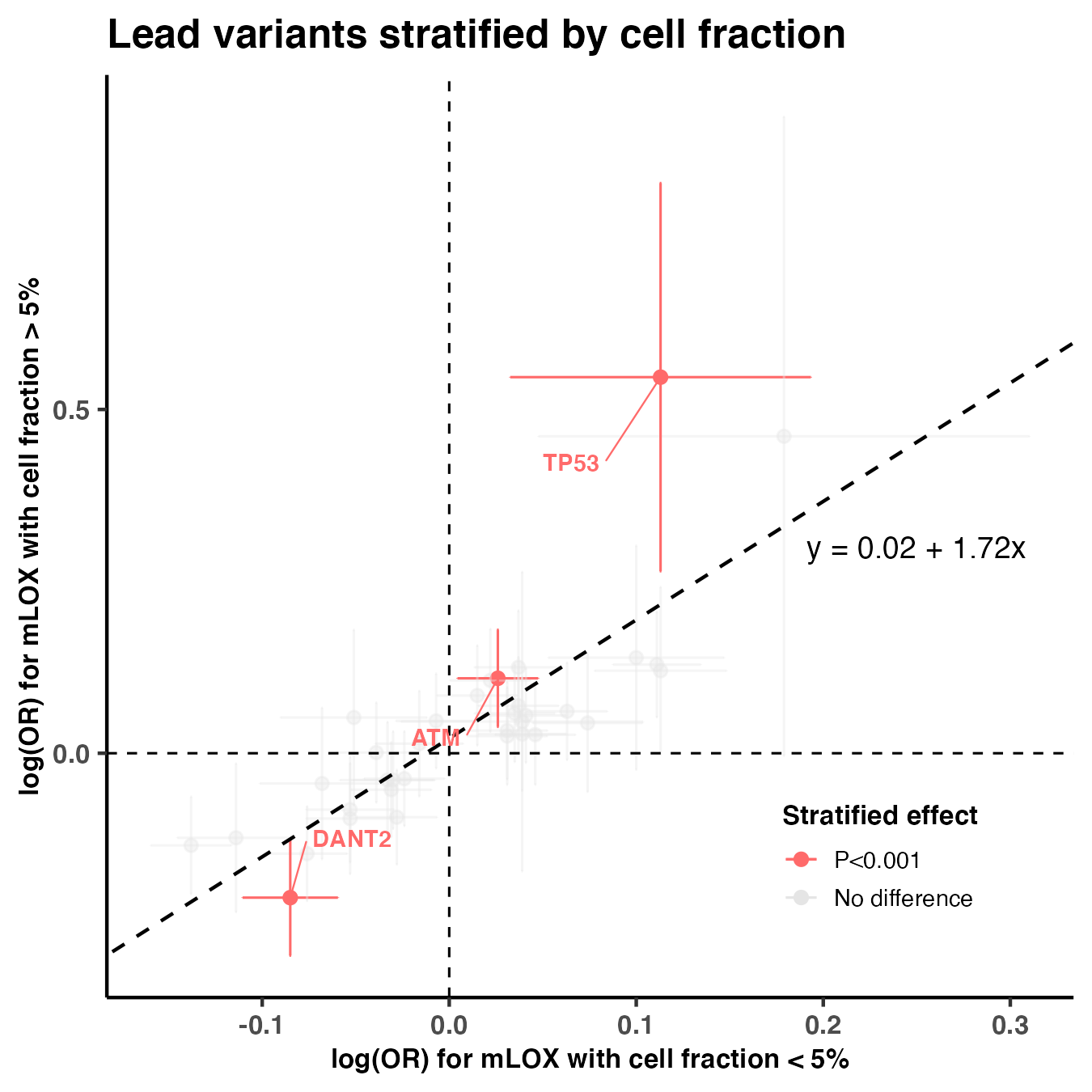

**Figure S8. Prevalence of different types of mosaic chromosomal alterations (mCA) by age at genotyping in the FinnGen cohort.**

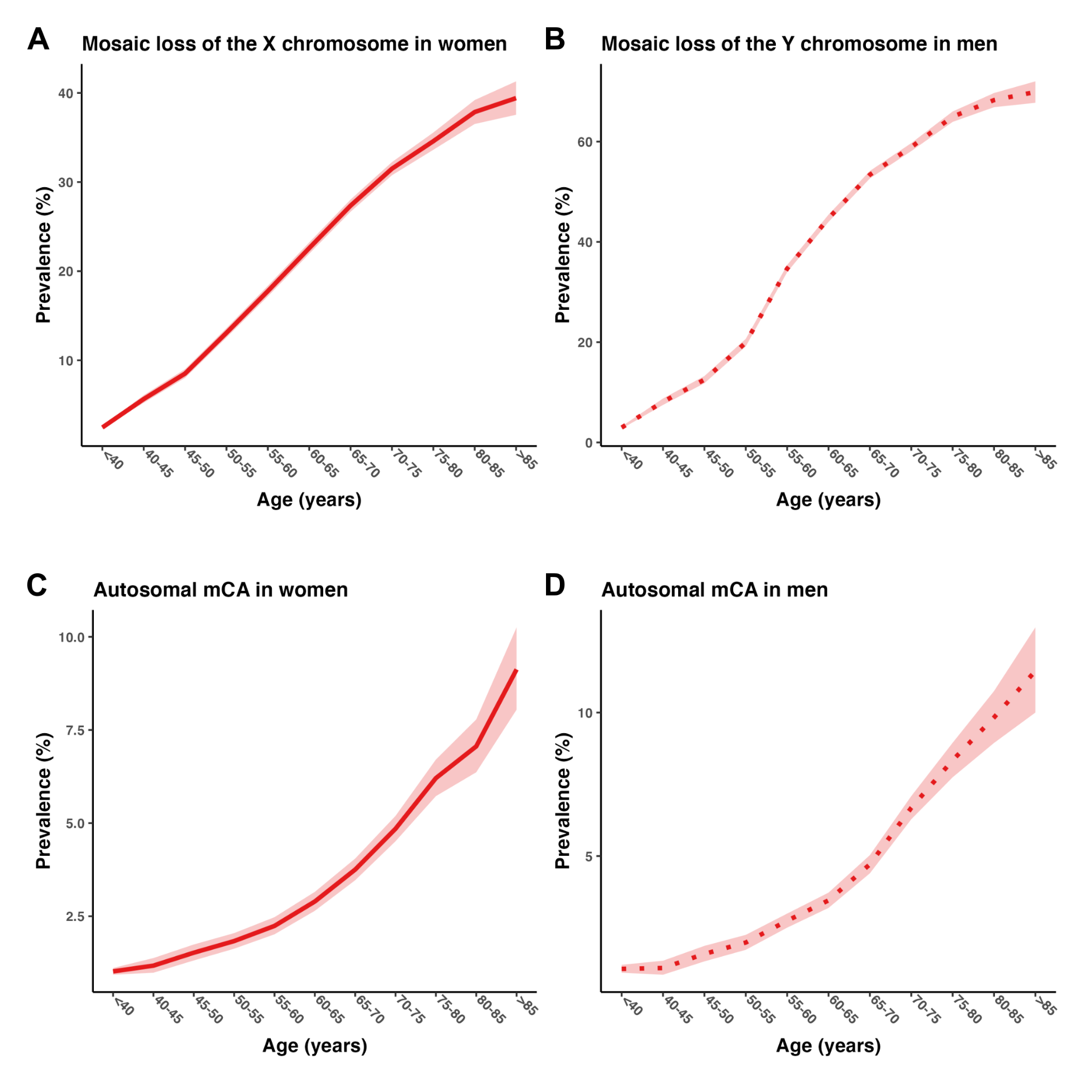

**Figure S9. Conditional GWAS adjusting for three HLA lead variants in the FinnGen cohort.**

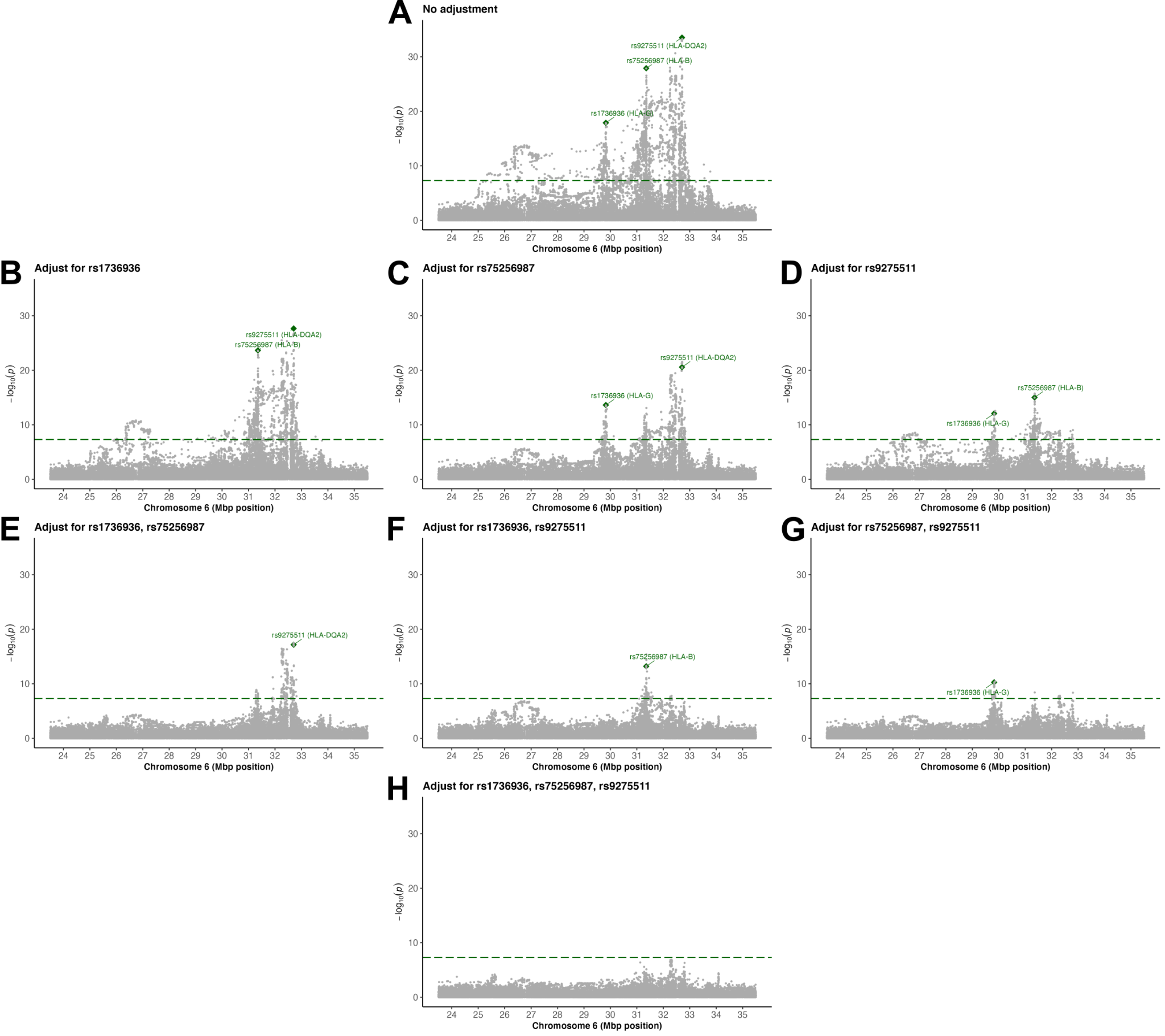

**Figure S10. The proportion of chromosome X variants being significant and the total number of variants examined in allelic shift analysis, with a 1k bp window.**

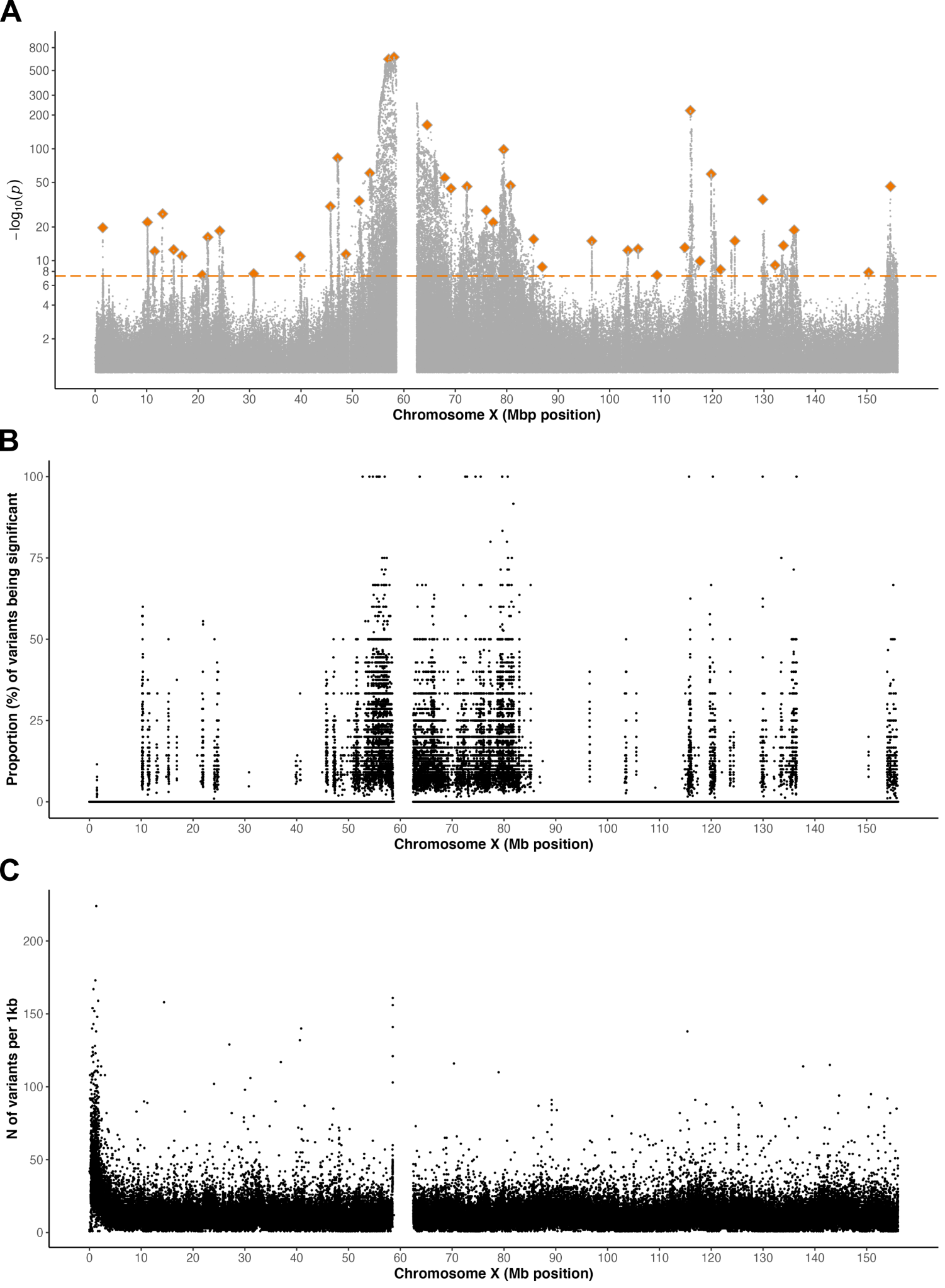

**Figure S11. Missense variants exhibiting significant allelic shifts in mLOX cases, with potential lead signals labeled.**

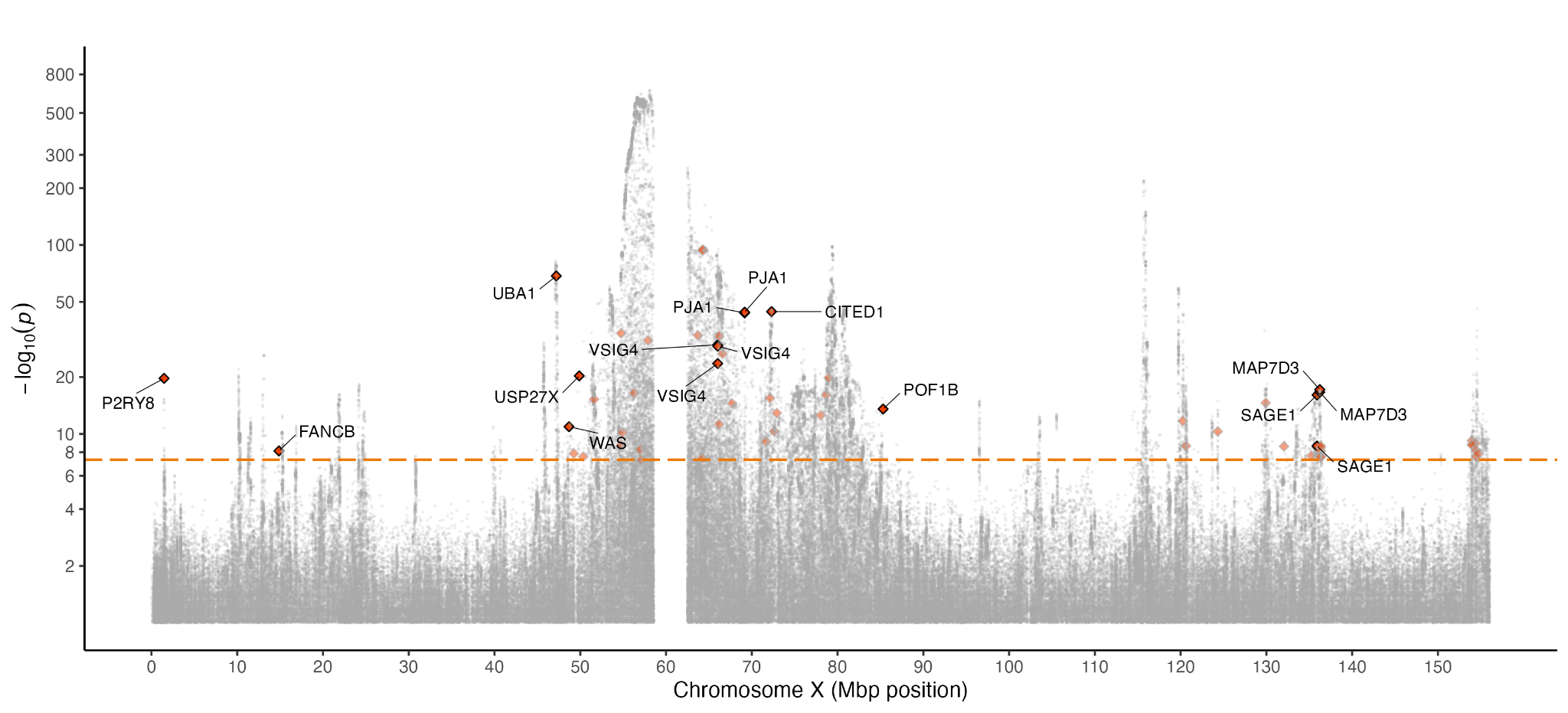

**Figure S12. Allelic shift in the context of X chromosome inactivation.**

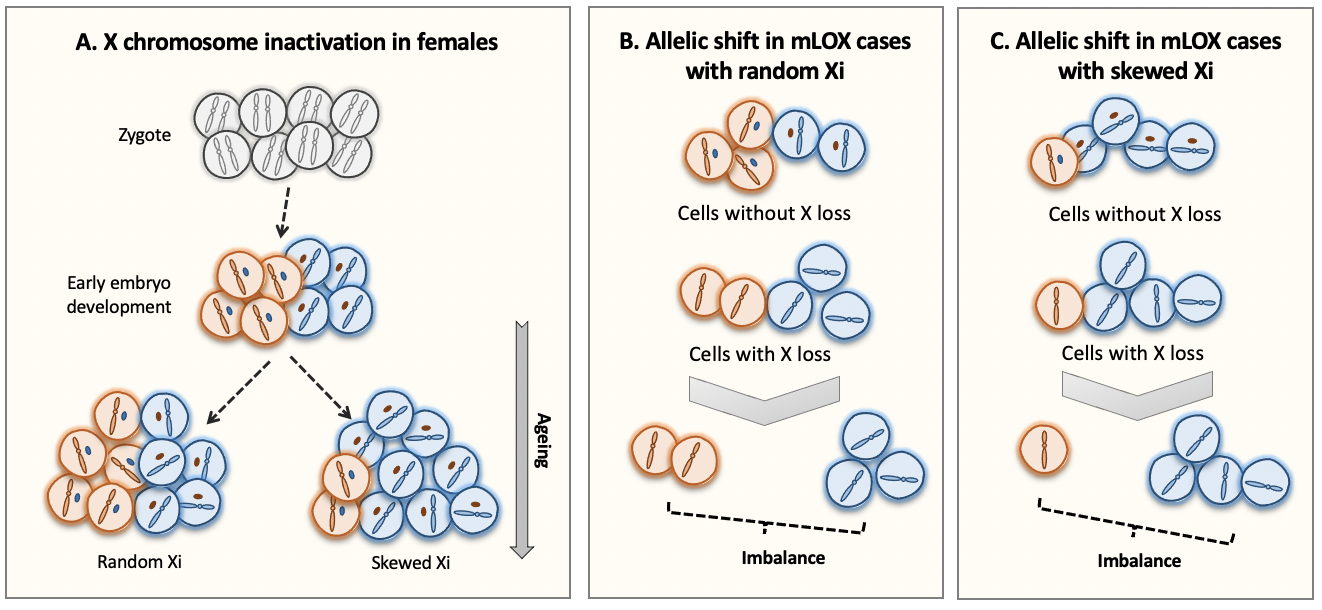

Panel (**A**) depicts the main mechanism of X chromosome inactivation (Xi) in females. To compensate for gene dosage imbalances between XX females and XY males, one of the two X chromosomes in females is randomly inactivated early in embryonic development and this inactivation status is passed down to daughter cells. As some females age, the expected 1:1 ratio of inactivated maternal to paternal X chromosome copies can become skewed, if cells harboring one of the active X chromosomes is more frequent than the other. Panel (**B**) and (**C**) depict the pattern of allelic shift in mLOX cases in terms of the status of Xi, with Panel (**B**) for random Xi and panel (**C**) for skewed Xi. As mLOX preferentially affects the inactivated X chromosome (Machiela et al., Nat Commun, 2016), the imbalance between chromosome X alleles in mLOX cases can be seen as the combined *cis* effects of both skewed Xi and mLOX. In other words, the imbalance of chromosome X alleles in mLOX cases could also been shaped by alleles that have *cis* effects solely on the process of skewed Xi.

**Figure S13. Prediction performance of X chromosome differential score constructed from 44 allelic shift analysis lead variants by score percentile, in real data (FinnGen) and simulated data.**

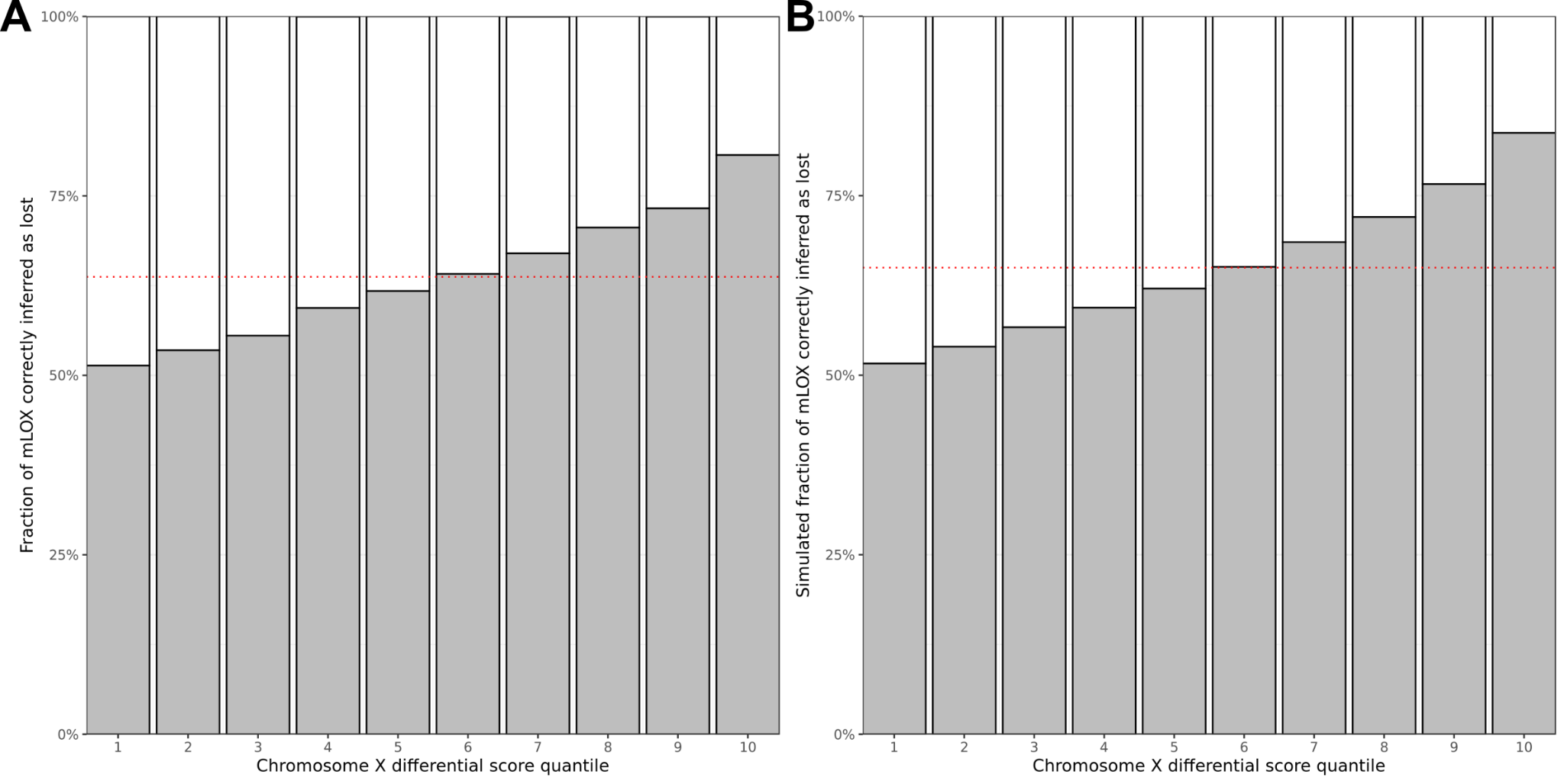

**Figure S14. Prediction performance contributed by each allelic shift analysis lead variant in real data (FinnGen) and simulated data.**

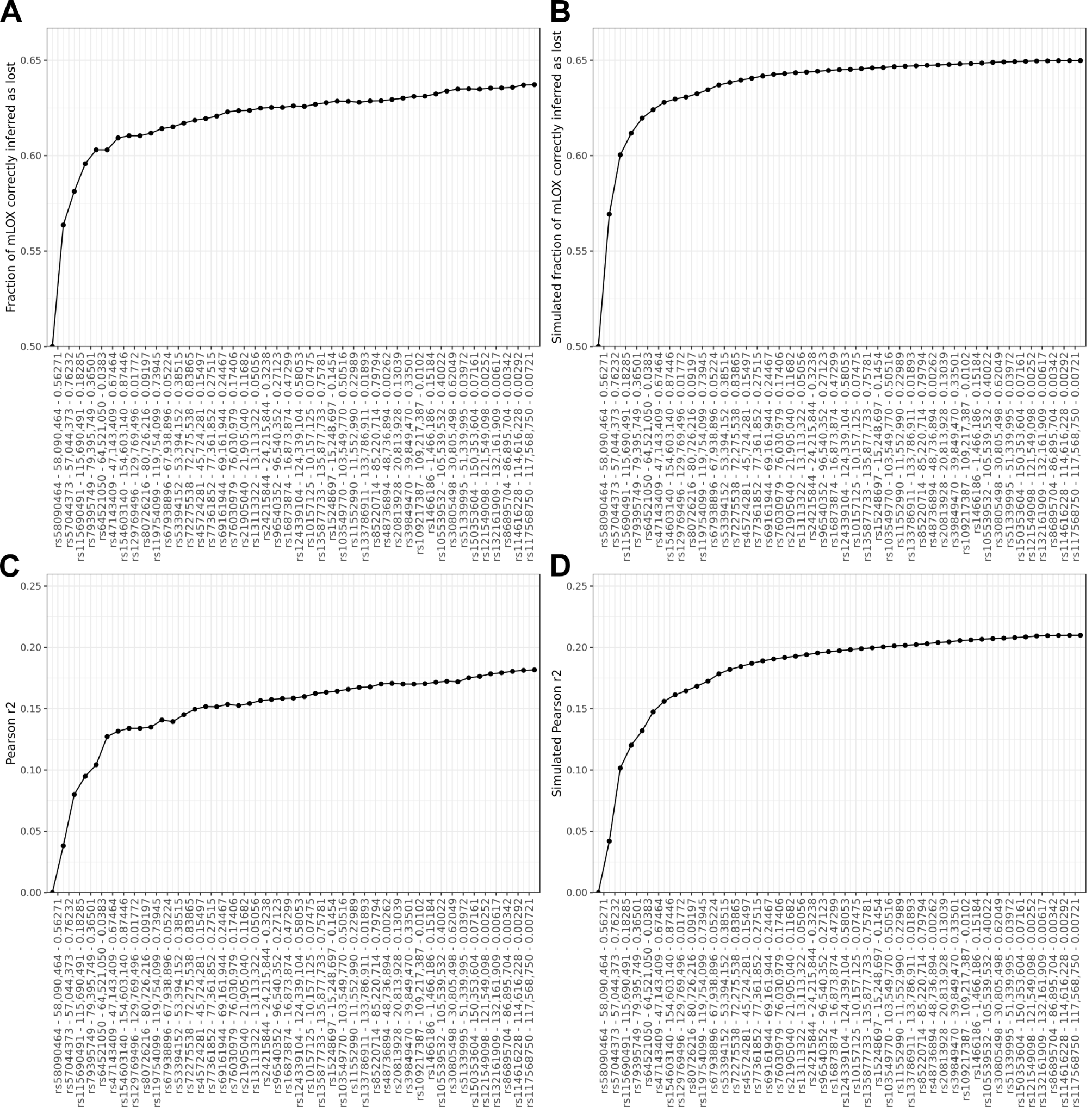

**Figure S15. The mechanism leading to detectable mLOX in women and mLOY in men.**

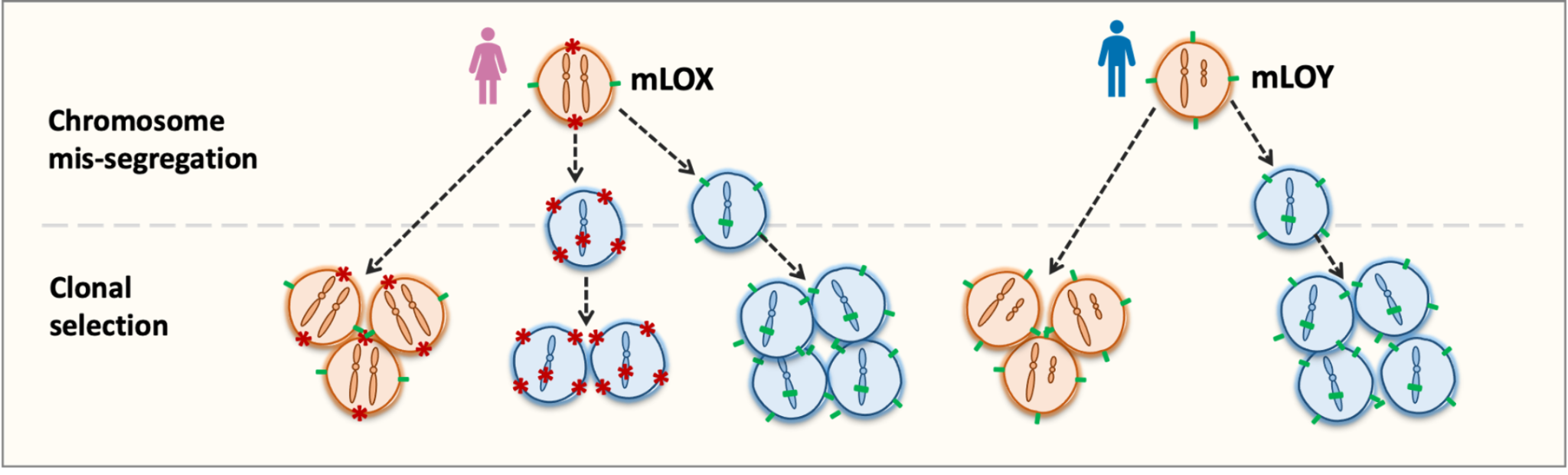

During mitosis, chromosome missegregation can cause the haematopoietic stem cells in females to lose one of the two X chromosomes (mLOX) or in males the Y chromosome (mLOY). For females, two lineages of mLOX mutant cells, presenting different X-linked antigens at the cell surface, are competing with each other during clonal expansion. Compared to mLOX in females, for males, the selection pressure for mLOY mutant cells could be weaker due to the limited number of Y antigens, and thus, the development of detectable mLOY is more impacted by the chromosome missegregation process.
