## Supplementary material for "Population analyses of mosaic X chromosome loss identify genetic drivers and widespread signatures of cellular selection": BCAC banner authorship

BCAC Authors

Thomas U. Ahearn^1^, Irene L. Andrulis^2, 3^, Hoda Anton-Culver^4^, Antonis C. Antoniou^5^, Amy Berrington^6^, Natalia V. Bogdanova^7-9^, Stig E. Bojesen^10-12^, Manjeet K. Bolla^5^, Hermann Brenner^13-15^, Jenny Chang-Claude^16, 17^, Georgia Chenevix-Trench^18^, Sarah V. Colonna^19^, CTS Consortium^20, 21^, Fergus J. Couch^22^, Angela Cox^23^, Kamila Czene^24^, Mary B. Daly^25^, Peter Devilee^26, 27^, Thilo Dörk^8^, Alison M. Dunning^28^, Miriam Dwek^29^, Douglas F. Easton^5, 28^, Diana M. Eccles^30^, Peter A. Fasching^31^, Olivia Fletcher^32^, Manuela Gago-Dominguez^33^, Montserrat García-Closas^1^, Mark S. Goldberg^34, 35^, Anna González-Neira^36^, Pascal Guénel^37^, Christopher A. Haiman^38^, Per Hall^24, 39^, Ute Hamann^40^, Antoinette Hollestelle^41^, Reiner Hoppe^42, 43^, John L. Hopper^44^, ABCTB Investigators^45^, Anna Jakubowska^46, 47^, Helena Jernström^48^, Esther M. John^49, 50^, Rudolf Kaaks^16^, Elza K. Khusnutdinova^51, 52^, Cari M. Kitahara^53^, Peter Kraft^54, 55^, Vessela N. Kristensen^56, 57^, Diether Lambrechts^58, 59^, Annika Lindblom^60, 61^, Arto Mannermaa^62-64^, Usha Menon^65^, Kyriaki Michailidou^5, 66^, Rachel A. Murphy^67, 68^, Heli Nevanlinna^69^, Nadia Obi^70^, Kenneth Offit^71, 72^, Paolo Peterlongo^73^, Paul D.P. Pharoah^5, 28, 74^, Dijana Plaseska-Karanfilska^75^, Gad Rennert^76^, Atocha Romero^77^, Emmanouil Saloustros^78^, Marjanka K. Schmidt^79-81^, Rita K. Schmutzler^82-84^, Jennifer Stone^44, 85^, Rulla M. Tamimi^55, 86^, Lauren R. Teras^87^, Mary Beth Terry^88^, Melissa A. Troester^89^, Celine M. Vachon^90^, Qin Wang^5^, Clarice R. Weinberg^91^, Robert Winqvist^92, 93^, Alicja Wolk^94^,

^1^ Division of Cancer Epidemiology and Genetics, National Cancer Institute, National Institutes of Health, Department of Health and Human Services, Bethesda, MD, USA.

^2^ Fred A. Litwin Center for Cancer Genetics, Lunenfeld-Tanenbaum Research Institute of Mount Sinai Hospital, Toronto, Ontario, Canada.

^3^ Department of Molecular Genetics, University of Toronto, Toronto, Ontario, Canada.

^4^ Department of Medicine, Genetic Epidemiology Research Institute, University of California Irvine, Irvine, CA, USA.

^5^ Centre for Cancer Genetic Epidemiology, Department of Public Health and Primary Care, University of Cambridge, Cambridge, UK.

^6^ Division of Genetics and Epidemiology, The Institute of Cancer Research, London, UK.

^7^ Department of Radiation Oncology, Hannover Medical School, Hannover, Germany.

^8^ Gynaecology Research Unit, Hannover Medical School, Hannover, Germany.

^9^ N.N. Alexandrov Research Institute of Oncology and Medical Radiology, Minsk, Belarus.

^10^ Copenhagen General Population Study, Herlev and Gentofte Hospital, Copenhagen University Hospital, Herlev, Denmark.

^11^ Department of Clinical Biochemistry, Herlev and Gentofte Hospital, Copenhagen University Hospital, Herlev, Denmark.

^12^ Faculty of Health and Medical Sciences, University of Copenhagen, Copenhagen, Denmark.

^13^ Division of Clinical Epidemiology and Aging Research, German Cancer Research Center (DKFZ), Heidelberg, Germany.

^14^ Division of Preventive Oncology, German Cancer Research Center (DKFZ) and National Center for Tumor Diseases (NCT), Heidelberg, Germany.

^15^ German Cancer Consortium (DKTK), German Cancer Research Center (DKFZ), Heidelberg, Germany.

^16^ Division of Cancer Epidemiology, German Cancer Research Center (DKFZ), Heidelberg, Germany.

^17^ Cancer Epidemiology Group, University Cancer Center Hamburg (UCCH), University Medical Center Hamburg-Eppendorf, Hamburg, Germany.

^18^ Cancer Division, QIMR Berghofer Medical Research Institute, Brisbane, Queensland, Australia.

^19^ Department of Internal Medicine and Huntsman Cancer Institute, University of Utah, Salt Lake City, UT, USA.

^20^ Department of Computational and Quantitative Medicine, City of Hope, Duarte, CA, USA.

^21^ City of Hope Comprehensive Cancer Center, City of Hope, Duarte, CA, USA.

^22^ Department of Laboratory Medicine and Pathology, Mayo Clinic, Rochester, MN, USA.

^23^ Sheffield Institute for Nucleic Acids (SInFoNiA), Department of Oncology and Metabolism, University of Sheffield, Sheffield, UK.

^24^ Department of Medical Epidemiology and Biostatistics, Karolinska Institutet, Stockholm, Sweden.

^25^ Department of Clinical Genetics, Fox Chase Cancer Center, Philadelphia, PA, USA.

^26^ Department of Pathology, Leiden University Medical Center, Leiden, the Netherlands.

^27^ Department of Human Genetics, Leiden University Medical Center, Leiden, the Netherlands.

^28^ Centre for Cancer Genetic Epidemiology, Department of Oncology, University of Cambridge, Cambridge, UK.

^29^ School of Life Sciences, University of Westminster, London, UK.

^30^ Faculty of Medicine, University of Southampton, Southampton, UK.

^31^ Department of Gynecology and Obstetrics, Comprehensive Cancer Center Erlangen-EMN, Friedrich-Alexander University Erlangen-Nuremberg, University Hospital Erlangen, Erlangen, Germany.

^32^ The Breast Cancer Now Toby Robins Research Centre, The Institute of Cancer Research, London, UK.

^33^ Genomic Medicine Group, International Cancer Genetics and Epidemiology Group, Fundación Pública Galega de Medicina Xenómica, Instituto de Investigación Sanitaria de Santiago de Compostela (IDIS), Complejo Hospitalario Universitario de Santiago, SERGAS, Santiago de Compostela, Spain.

^34^ Department of Medicine, McGill University, Montréal, QC, Canada.

^35^ Division of Clinical Epidemiology, Royal Victoria Hospital, McGill University, Montréal, QC, Canada.

^36^ Human Genotyping Unit-CeGen, Spanish National Cancer Research Centre (CNIO), Madrid, Spain.

^37^ Team Exposome and Heredity, CESP, Gustave Roussy, INSERM, University Paris-Saclay, UVSQ, Villejuif, France.

^38^ Department of Preventive Medicine, Keck School of Medicine, University of Southern California, Los Angeles, CA, USA.

^39^ Department of Oncology, Södersjukhuset, Stockholm, Sweden.

^40^ Molecular Genetics of Breast Cancer, German Cancer Research Center (DKFZ), Heidelberg, Germany.

^41^ Department of Medical Oncology, Erasmus MC Cancer Institute, Rotterdam, the Netherlands.

^42^ Dr. Margarete Fischer-Bosch-Institute of Clinical Pharmacology, Stuttgart, Germany.

^43^ University of Tübingen, Tübingen, Germany.

^44^ Centre for Epidemiology and Biostatistics, Melbourne School of Population and Global Health, The University of Melbourne, Melbourne, Victoria, Australia.

^45^ Australian Breast Cancer Tissue Bank, Westmead Institute for Medical Research, University of Sydney, Sydney, New South Wales, Australia.

^46^ Department of Genetics and Pathology, International Hereditary Cancer Center, Pomeranian Medical University, Szczecin, Poland.

^47^ Independent Laboratory of Molecular Biology and Genetic Diagnostics, Pomeranian Medical University, Szczecin, Poland.

^48^ Oncology, Department of Clinical Sciences in Lund, Lund University, Lund, Sweden.

^49^ Department of Epidemiology and Population Health, Stanford University School of Medicine, Stanford, CA, USA.

^50^ Department of Medicine, Division of Oncology, Stanford Cancer Institute, Stanford University School of Medicine, Stanford, CA, USA.

^51^ Institute of Biochemistry and Genetics of the Ufa Federal Research Centre of the Russian Academy of Sciences, Ufa, Russia.

^52^ Department of Genetics and Fundamental Medicine, Bashkir State University, Ufa, Russia.

^53^ Radiation Epidemiology Branch, Division of Cancer Epidemiology and Genetics, National Cancer Institute, Bethesda, MD, USA.

^54^ Program in Genetic Epidemiology and Statistical Genetics, Harvard T.H. Chan School of Public Health, Boston, MA, USA.

^55^ Department of Epidemiology, Harvard T.H. Chan School of Public Health, Boston, MA, USA.

^56^ Department of Medical Genetics, Oslo University Hospital and University of Oslo, Oslo, Norway.

^57^ Institute of Clinical Medicine, Faculty of Medicine, University of Oslo, Oslo, Norway.

^58^ Laboratory for Translational Genetics, Department of Human Genetics, KU Leuven, Leuven, Belgium.

^59^ VIB Center for Cancer Biology, VIB, Leuven, Belgium.

^60^ Department of Molecular Medicine and Surgery, Karolinska Institutet, Stockholm, Sweden.

^61^ Department of Clinical Genetics, Karolinska University Hospital, Stockholm, Sweden.

^62^ Translational Cancer Research Area, University of Eastern Finland, Kuopio, Finland.

^63^ Institute of Clinical Medicine, Pathology and Forensic Medicine, University of Eastern Finland, Kuopio, Finland.

^64^ Biobank of Eastern Finland, Kuopio University Hospital, Kuopio, Finland.

^65^ MRC Clinical Trials Unit, Institute of Clinical Trials & Methodology, University College London, London, UK.

^66^ Biostatistics Unit, The Cyprus Institute of Neurology & Genetics, Nicosia, Cyprus.

^67^ School of Population and Public Health, University of British Columbia, Vancouver, BC, Canada.

^68^ Cancer Control Research, BC Cancer Agency, Vancouver, BC, Canada.

^69^ Department of Obstetrics and Gynecology, Helsinki University Hospital, University of Helsinki, Helsinki, Finland.

^70^ Institute for Medical Biometry and Epidemiology, University Medical Center Hamburg-Eppendorf, Hamburg, Germany.

^71^ Clinical Genetics Research Lab, Department of Cancer Biology and Genetics, Memorial Sloan Kettering Cancer Center, New York, NY, USA.

^72^ Clinical Genetics Service, Department of Medicine, Memorial Sloan Kettering Cancer Center, New York, NY, USA.

^73^ Genome Diagnostics Program, IFOM ETS - the AIRC Institute of Molecular Oncology, Milan, Italy.

^74^ Department of Computational Biomedicine, Cedars-Sinai Medical Center, West Hollywood, CA, USA.

^75^ Research Centre for Genetic Engineering and Biotechnology Georgi D. Efremov, MASA, Skopje, Republic of North Macedonia.

^76^ Clalit National Cancer Control Center, Carmel Medical Center and Technion Faculty of Medicine, Haifa, Israel.

^77^ Medical Oncology Department, Hospital Universitario Puerta de Hierro, Madrid, Spain.

^78^ Department of Oncology, University Hospital of Larissa, Larissa, Greece.

^79^ Division of Molecular Pathology, The Netherlands Cancer Institute, Amsterdam, the Netherlands.

^80^ Division of Psychosocial Research and Epidemiology, The Netherlands Cancer Institute - Antoni van Leeuwenhoek hospital, Amsterdam, the Netherlands.

^81^ Department of Clinical Genetics, Leiden University Medical Center, Leiden, the Netherlands.

^82^ Center for Familial Breast and Ovarian Cancer, Faculty of Medicine and University Hospital Cologne, University of Cologne, Cologne, Germany.

^83^ Center for Integrated Oncology (CIO), Faculty of Medicine and University Hospital Cologne, University of Cologne, Cologne, Germany.

^84^ Center for Molecular Medicine Cologne (CMMC), Faculty of Medicine and University Hospital Cologne, University of Cologne, Cologne, Germany.

^85^ Genetic Epidemiology Group, School of Population and Global Health, University of Western Australia, Perth, Western Australia, Australia.

^86^ Department of Population Health Sciences, Weill Cornell Medicine, New York, NY, USA.

^87^ Department of Population Science, American Cancer Society, Atlanta, GA, USA.

^88^ Department of Epidemiology, Mailman School of Public Health, Columbia University, New York, NY, USA.

^89^ Department of Epidemiology, Gillings School of Global Public Health and UNC Lineberger Comprehensive Cancer Center, University of North Carolina at Chapel Hill, Chapel Hill, NC, USA.

^90^ Department of Quantitative Health Sciences, Division of Epidemiology, Mayo Clinic, Rochester, MN, USA.

^91^ Biostatistics and Computational Biology Branch, National Institute of Environmental Health Sciences, NIH, Research Triangle Park, NC, USA.

^92^ Laboratory of Cancer Genetics and Tumor Biology, Translational Medicine Research Unit, Biocenter Oulu, University of Oulu, Oulu, Finland.

^93^ Laboratory of Cancer Genetics and Tumor Biology, Northern Finland Laboratory Centre Oulu, Oulu, Finland.

^94^ Institute of Environmental Medicine, Karolinska Institutet, Stockholm, Sweden.
