## Supplementary material for "Population analyses of mosaic X chromosome loss identify genetic drivers and widespread signatures of cellular selection": MVP banner authorship

**Million Veteran Program: Consortium Acknowledgement for Manuscripts**

**MVP Executive Committee**

- Co-Chair: J. Michael Gaziano, M.D., M.P.H.

- Co-Chair: Rachel Ramoni, D.M.D., Sc.D.

- Jim Breeling, M.D. (ex-officio)

- Kyong-Mi Chang, M.D.

- Grant Huang, Ph.D.

- Sumitra Muralidhar, Ph.D.

- Christopher J. O’Donnell, M.D., M.P.H.

- Philip S. Tsao, Ph.D.

**MVP Program Office**

- Sumitra Muralidhar, Ph.D.

- Jennifer Moser, Ph.D.

**MVP Recruitment/Enrollment**

- Recruitment/Enrollment Director/Deputy Director, Boston

- Stacey B. Whitbourne, Ph.D.; Jessica V. Brewer, M.P.H.

- MVP Coordinating Centers

o Clinical Epidemiology Research Center (CERC), West Haven – John Concato, M.D., M.P.H.

o Cooperative Studies Program Clinical Research Pharmacy Coordinating Center, Albuquerque - Stuart Warren, J.D., Pharm D.; Dean P. Argyres, M.S.

o Genomics Coordinating Center, Palo Alto – Philip S. Tsao, Ph.D.

o Massachusetts Veterans Epidemiology Research Information Center (MAVERIC), Boston - J. Michael Gaziano, M.D., M.P.H.

o MVP Information Center, Canandaigua – Brady Stephens, M.S.

- Core Biorepository, Boston – Mary T. Brophy M.D., M.P.H.; Donald E. Humphries, Ph.D.

- MVP Informatics, Boston – Nhan Do, M.D.; Shahpoor Shayan

- Data Operations/Analytics, Boston – Xuan-Mai T. Nguyen, Ph.D.

**MVP Science**

- Genomics - Christopher J. O’Donnell, M.D., M.P.H.; Saiju Pyarajan Ph.D.; Philip S. Tsao, Ph.D.

- Phenomics - Kelly Cho, M.P.H, Ph.D.

- Data and Computational Sciences – Saiju Pyarajan, Ph.D.

- Statistical Genetics – Elizabeth Hauser, Ph.D.; Yan Sun, Ph.D.; Hongyu Zhao, Ph.D.

**MVP Local Site Investigators**

- Atlanta VA Medical Center (Peter Wilson) - Bay Pines VA Healthcare System (Rachel McArdle)

- Birmingham VA Medical Center (Louis Dellitalia)

- Cincinnati VA Medical Center (John Harley)

- Clement J. Zablocki VA Medical Center (Jeffrey Whittle)

- Durham VA Medical Center (Jean Beckham)

- Edith Nourse Rogers Memorial Veterans Hospital (John Wells)

- Edward Hines, Jr. VA Medical Center (Salvador Gutierrez)

- Fayetteville VA Medical Center (Gretchen Gibson)

- VA Health Care Upstate New York (Laurence Kaminsky)

- New Mexico VA Health Care System (Gerardo Villareal)

- VA Boston Healthcare System (Scott Kinlay)

- VA Western New York Healthcare System (Junzhe Xu)

- Ralph H. Johnson VA Medical Center (Mark Hamner)

- Wm. Jennings Bryan Dorn VA Medical Center (Kathlyn Sue Haddock)

- VA North Texas Health Care System (Sujata Bhushan)

- Hampton VA Medical Center (Pran Iruvanti)

- Hunter Holmes McGuire VA Medical Center (Michael Godschalk)

- Iowa City VA Health Care System (Zuhair Ballas)

- Jack C. Montgomery VA Medical Center (Malcolm Buford)

- James A. Haley Veterans’ Hospital (Stephen Mastorides)

- Louisville VA Medical Center (Jon Klein)

- Manchester VA Medical Center (Nora Ratcliffe)

- Miami VA Health Care System (Hermes Florez)

- Michael E. DeBakey VA Medical Center (Alan Swann)

- Minneapolis VA Health Care System (Maureen Murdoch)

- N. FL/S. GA Veterans Health System (Peruvemba Sriram)

- Northport VA Medical Center (Shing Shing Yeh)

- Overton Brooks VA Medical Center (Ronald Washburn)

- Philadelphia VA Medical Center (Darshana Jhala)

- Phoenix VA Health Care System (Samuel Aguayo)

- Portland VA Medical Center (David Cohen)

- Providence VA Medical Center (Satish Sharma)

- Richard Roudebush VA Medical Center (John Callaghan)

- Salem VA Medical Center (Kris Ann Oursler)

- San Francisco VA Health Care System (Mary Whooley)

- South Texas Veterans Health Care System (Sunil Ahuja)

- Southeast Louisiana Veterans Health Care System (Amparo Gutierrez)

- Southern Arizona VA Health Care System (Ronald Schifman)

- Sioux Falls VA Health Care System (Jennifer Greco)

- St. Louis VA Health Care System (Michael Rauchman)

- Syracuse VA Medical Center (Richard Servatius)

- VA Eastern Kansas Health Care System (Mary Oehlert)

- VA Greater Los Angeles Health Care System (Agnes Wallbom)

- VA Loma Linda Healthcare System (Ronald Fernando)

- VA Long Beach Healthcare System (Timothy Morgan)

- VA Maine Healthcare System (Todd Stapley)

- VA New York Harbor Healthcare System (Scott Sherman)

- VA Pacific Islands Health Care System (Gwenevere Anderson)

- VA Palo Alto Health Care System (Philip Tsao)

- VA Pittsburgh Health Care System (Elif Sonel)

- VA Puget Sound Health Care System (Edward Boyko)

- VA Salt Lake City Health Care System (Laurence Meyer)

- VA San Diego Healthcare System (Samir Gupta)

- VA Southern Nevada Healthcare System (Joseph Fayad)

- VA Tennessee Valley Healthcare System (Adriana Hung)

- Washington DC VA Medical Center (Jack Lichy)

- W.G. (Bill) Hefner VA Medical Center (Robin Hurley)

- White River Junction VA Medical Center (Brooks Robey)

- William S. Middleton Memorial Veterans Hospital (Robert Striker)
